# Transdiagnostic quantitative assessment of dementias using in vivo MRI and data-driven disease progression modelling: a case study in Alzheimer’s disease and dementia with Lewy bodies

**DOI:** 10.1101/2025.10.03.25337171

**Authors:** Gonzalo Castro Leal, Ajay Konuri, Alexandra L. Young, Niloufar Zebarjadi, Tanmayee Samantaray, Annegret Habich, Nicolás Castellanos-Perilla, María Camila Gonzalez, John-Paul Taylor, Michael Firbank, Alan Thomas, Paul C Donaghy, John T O’Brien, Simon J.G. Lewis, Daniel Alcolea, Alexandre Bejanin, Andrea Pilotto, Alessandro Padovani, Afina W. Lemstra, Mara ten Kate, Kurt Segers, Irena Rektorova, Ahmet Turan Isik, Bedia Samanci, Consuelo Cháfer-Pericás, Christian Lambert, Ramón Landin-Romero, Rohan Bhome, Ivelina Dobreva, Rimona S. Weil, Zuzana Walker, Dag Aarsland, Eric Westman, Daniel Ferreira, Elie Matar, Neil P. Oxtoby, the Alzheimer’s Disease Neuroimaging Initiative

## Abstract

**INTRODUCTION:** Alzheimer’s disease (AD) and Lewy body disease cause brain atrophy producing dementia syndromes with shared clinical symptoms. We hypothesise that those diseases present with differential atrophy patterns, thus making disease progression modelling methods useful for biological staging.

**METHODS:** Using features derived from magnetic resonance imaging (MRI) scans from six international cohorts, we applied disease progression subtype and stage modelling on brain volume data from patients across Alzheimer’s disease dementia (ADD) and dementia with Lewy bodies (DLB) diagnoses. We assessed clinical, biomarker, and histopathological associations with the discovered atrophy subtypes.

**RESULTS:** We identified three data-driven brain atrophy subtypes across syndromes that could support biologically informed diagnosis in vivo: Limbic, Cortico-Limbic, and Cortical. Clinical syndrome and post-mortem assessments aligned imperfectly, but plausibly, with subtype: Limbic (more AD), Cortical (more DLB), Cortico-Limbic (mixed). In particular, the Limbic and Cortico-Limbic subtypes showed higher amyloid and tau positivity, and worse memory impairment than the Cortical subtype.

**CONCLUSION:** Our novel data-driven transdiagnostic approach shows promise for supporting in vivo biological diagnosis and subtyping of patients using only a single-visit MRI.

## Introduction

Two common dementia syndromes in older adults are (probable) Alzheimer’s disease dementia (ADD) and dementia with Lewy bodies (DLB). Differential diagnosis is challenging due to significant overlap in the cognitive and neuropsychological presentations, especially at early stages where symptoms are subtle.^1–3^

Each syndrome is primarily caused by a specific neurodegenerative disease: Alzheimer’s disease (AD) and Lewy body disease (LBD), respectively. AD is defined, originally *post mortem* but recently also *in vivo* using biomarkers, by the presence in the brain of extracellular amyloid-beta (Aβ) plaques^4,5^ and intracellular neurofibrillary tangles (NFTs).^6,7^ LBD, which includes DLB among other syndromes, is pathologically characterised by inclusions of α-synuclein (α-syn) called Lewy bodies and Lewy neurites.^8^ Confirmation of these hallmarks during *post mortem* examination represents the gold standard for diagnosis of each disease.^9^

*In vivo* measurements of disease-specific pathology can be achieved through the detection of related metabolites in cerebrospinal fluid (CSF) and blood plasma samples,^10–16^ and/or in positron emission tomography (PET) imaging.^17–19^ Integration of these methods into diagnostic workflows has shifted ADD diagnosis in research studies from a symptom-based to a biological-based paradigm.^20^ However, clinical rollout of CSF and PET remains limited due to high cost/burden for healthcare systems and patients, while blood plasma methods have not yet been approved for clinical use.^21^ This has created an opportunity to develop new methods that leverage existing, routinely acquired data to identify and characterise underlying biological processes, potentially serving as companion diagnostics in the future.

Previous studies have explored the utility of routinely acquired structural magnetic resonance imaging (MRI) for differential diagnosis, particularly region-specific differences in brain volume at the group level.^22–25^ However, consensus is limited as to which regional volumes can differentiate dementias, possibly because many studies have ignored biological and phenotypic heterogeneity by grouping all patients together which can dilute disease signal (reduce statistical power).^26^ Several studies have attempted to explore this heterogeneity using clustering/subtyping. In ADD, consistent subtypes have emerged: Typical AD, Limbic predominant, Hippocampal-sparing, and Minimal atrophy.^27^ Studies in DLB are fewer, with one study identifying distinct clusters within DLB primarily driven by grey matter volume changes in the pallidum, caudate, cingulate, and olfactory cortex.^28^ Only one existing clustering approach, the Subtype and Stage Inference (SuStaIn) algorithm,^29^ is *progression aware, i.e.,* avoids confounding disease severity with disease phenotype. SuStaIn has been applied widely to characterise heterogeneity in neurodegenerative diseases.^29–33^

The known co-occurrence of AD and LBD copathology^34^ motivates exploration of advanced clustering methods across syndromes – that is, transdiagnostically – rather than separately. In this study, SuStaIn is leveraged to study MRI atrophy patterns of ADD and DLB patients to build the first transdiagnostic data-driven subtyping model of dementias. The model is assessed for utility across clinical, biological, and histopathological measures of interest.

## Methods

### Participants

We combined participant data from multiple cohorts: Alzheimer’s Disease Neuroimaging Initiative (ADNI), National Alzheimer’s Coordinating Center (NACC), European DLB Consortium (E-DLB), Parkinson’s Disease Biomarkers Program (PDBP), Vision in Parkinson’s Disease (ViPD, University College London), and Predicting Parkinson’s and Dementia (Pre-D, University of Sydney).

Data used in the preparation of this article were obtained from the Alzheimer’s Disease Neuroimaging Initiative (ADNI) database (http://adni.loni.usc.edu). The ADNI was launched in 2003 as a public-private partnership, led by Principal Investigator Michael W. Weiner, MD. The primary goal of ADNI has been to test whether serial magnetic resonance imaging (MRI), positron emission tomography (PET), other biological markers, and clinical and neuropsychological assessment can be combined to measure the progression of mild cognitive impairment (MCI) and early Alzheimer’s disease (AD).

Inclusion criteria for our study: clinical diagnosis of ADD, DLB, or MCI with suspected AD or LBD (see Supplementary Information: Diagnostic criteria); and a T1-weighted (T1w) brain MRI scan. Normative data was sourced from cognitively normal (CN) participants (see Transdiagnostic Subtyping Framework). All data were collected with appropriate informed consent under ethically approved clinical studies. Our secondary analysis of these data was approved by the UCL Research Ethics Committee under application 8019/005 and the UCL Department of Computer Science Research Ethics Committee under application UCL-CSREC-209-B.

### Participant Data

Data availability varied across cohorts (see **Table S2** for a detailed breakdown). The input features to our transdiagnostic subtype model were derived from T1w MRI (see Imaging Derived Phenotypes), adjusted for demographics (sex, age) and total intracranial volume. Our *post-hoc* analysis (see Statistical Analysis) compared and contrasted subtypes using cohort-specific clinical, biomarker, genetic, and neuropathological features.

Clinical measures included Mini Mental State Examination (MMSE) and/or Montreal Cognitive Assessment (MoCA) scores as well as domain-specific scores for memory, language, executive, and visuospatial cognition from the Alzheimer’s Disease Sequencing Project Phenotype Harmonization Consortium (ADSP-PHC)^35^. DLB cohorts included measures of core clinical features of DLB (hallucinations, REM sleep behaviour disorder (RBD), cognitive fluctuations, and parkinsonism). In E-DLB, many of the patients were assessed before the 2017 International Consensus Criteria^1^, so the presence/absence of clinical features was based on the 2005 International Consensus Criteria^36^ for probable DLB to harmonize diagnosis across centres.

Biomarker data included CSF measurements of Amyloid-beta (Aβ) 1-42, Phospho-Tau (p-tau) 181 (Elecsys),^37^ and α-syn analysed using Seed Amplification Assay (SAA),^38^ with data excluded if beyond 200 to 1700 pg/mL for Aβ and 8 to 120 pg/mL for p-tau. Imaging data included Aβ PET from multiple tracers (centiloid) and tau PET (18F-AV-1451) standardised uptake value ratio (SUVR) at the lobe level. Genetics analysis was limited to Apolipoprotein E ε4 (*APOE* ε4) allele number of copies.

Neuropathological data included AD and LBD captured in the format of the Neuropathology Data Form of NACC. AD neuropathological confirmation (ADNC) was reported as none, low, mid or high, depending on the presence and location of amyloid plaques, NFTs, and neuritic plaques.^39^ Lewy body presence was reported as none, diffuse, brainstem-, limbic-, or neocortical-predominant.

### Imaging Derived Phenotypes

We processed structural T1-weighted MPRAGE-like MRI scans with the FreeSurfer image analysis suite (https://surfer.nmr.mgh.harvard.edu/) to produce regional brain tissue volumes using the Desikan-Killiany atlas parcellation. Technical details are described in prior publications.^40–49^ We used FreeSurfer version 7.1.1 except for E-DLB scans, which were processed with FreeSurfer version 7.3.2 (compatible with 7.1.1). E-DLB data was pre-processed and stored at the E-DLB MRI Hub at Karolinska Institute (Stockholm) within the HiveDB.^50^

The brain volumes were candidate inputs to our transdiagnostic model, with bilateral subcortical and cortical lobe regions of interest (ROIs) considered — see

**Table S1**. Inclusion of ROIs required a previously reported statistically significant difference in either syndrome (controls vs cases). Patient and MCI data was detrended (residuals method) for age, sex, and intracranial volume (ICV) and normalised (robust z-scores) using data from our reference population of cognitively normal participants having continuously confirmed normal cognition for a minimum of 3 years.

### Transdiagnostic Subtyping Framework

Our approach fits off-the-shelf SuStaIn models in each dementia syndrome separately, refining each model assignments using a Null Subtype approach (described below), then combines all subtypes into a single transdiagnostic model after removing redundant subtypes based on model similarity (Hellinger Distance)^51^ and cross-subtype explainability (R2 score between data likelihood of subtype pairs). The number of subtypes per syndrome is informed by the 10-fold cross-validation information criteria (CVIC), as is typical for SuStaIn analyses.^29^

SuStaIn discovers progression-aware clusters in the data, which are subtypes having distinct spatiotemporal signatures of regional brain tissue loss. Progression is modelled as piecewise linear accumulation of atrophy severity between z-score ‘events’, which we set to be z = 0.67, 1, 2 (75th, 86th, 98th percentiles of normative atrophy) to capture mid-to-late atrophy. SuStaIn simultaneously optimizes the subtype clusters and their event-based disease progression profiles. The likelihood equation to optimize is:

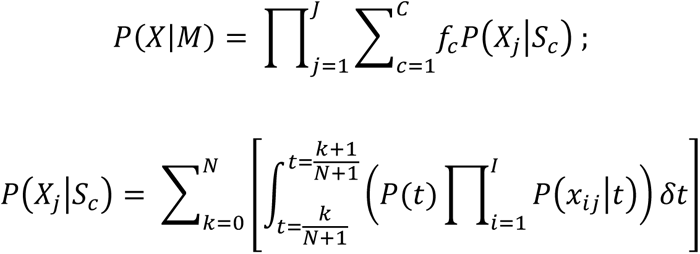

Where ***X*** = {*x_ij_* | *i* = 1, …, *I*; *j* = 1, …, *J*} is a set of measurements ***i*** for subject ***j***, ***M*** is the overall SuStaIn model, ***C*** is the number of clusters, ***f_c_*** is the fraction of subjects assigned to a particular subtype, ***N*** is the number of z-score events. The predefined set of input features (spatial dimension) and set of events (temporal dimension) defines the number of model stages, and the number of stages by subtypes plus the fraction parameter defines the total number of model parameters. In practice, spatial and temporal resolution of the model is constrained by computational complexity and model redundancy which must both be balanced with the available data. Our rule of thumb for a trustworthy model is a minimum ratio of 5:1 sample to parameters.

SuStaIn assigns data samples (individuals) to both subtype and stage via a likelihood normalized score (subtype probability).^29^ After model fitting, those in stage zero (of any subtype) tend to have sub-threshold abnormality and are re-assigned to a separate subgroup. However, during fitting of off-the-shelf SuStaIn, all data is forced into one of the clusters. This includes any outliers which, if present, would contaminate the progression pattern in each subtype/cluster. This also affects the model assignments of new data. Unfortunately, because SuStaIn output probability is calculated as a normalized likelihood ratio across subtypes per subject, rejecting assignments based on subtype probability would not discard weak subtype assignments – a subject’s data might fit poorly every sequence, but still produce an elevated ratio.

We introduce a Null Subtype to produce subtype assignments that are not contaminated by outliers. The Null Subtype consists of an artificial progression pattern whereby each z-score event occurs simultaneously across input features. This mimics an uninformative uniform distribution aimed at capturing all outliers. For full details, see Supplementary Information: The Null Subtype: Implementation and Implications. Our statistical comparison of subtypes (described below) excludes participant data assigned to the Null subtype and the Stage Zero subgroup. Since being assigned to the Null can be a consequence of either noise or not adhering any of the estimated patterns of atrophy, we perform the post-hoc analysis on both the first available visit (main analysis) and the first non-Null visit (extended analysis) which accounts for “noisy visits”.

### Statistical Analysis

All statistical analysis and visualizations were conducted using Python version 3.9. We performed subtype (and stage) associations over available clinical and biological markers of interest, using robust linear regression models. Some variables were transformed to reduce skew: MMSE transformed to the square root of the number of errors as described in Jacqmin-Gadda et al,^52^ and CSF measurements are log-transformed. Predefined cutoffs for the CSF p-tau/Aβ ratio were used to determine amyloid and tau positivity.^37,53^ Covariates included age, sex, educational level, and *APOE* ε4 when appropriate. Group level differences were explored using either Kruskal-Wallis or Mann-Whitney U test (in pairwise comparisons) for continuous data, and χ^2^ contingency test for categorical data. Effect sizes were reported as Cliff’s delta or Cramer’s V and residuals, respectively. We performed false discovery rate (FDR) adjustments for multiple comparisons, with the statistical significance after correction at p_FDR_ < 0.05.

## Results

### Participant characteristics

Demographic summaries of each cohort are presented in **Table 1**. The patient population was aged 72.5 ± 8.4 years (mean ± standard deviation), and 50.5% were male. Years of education (2.6% missing) was 15.4 ± 3.5 years, with 77.8% of patients having at least high school education. Cognitive scores were available for 85.8% of patients: MMSE in 58.6% of these, MoCA in 47.5% (20.4% had both).

ADNI and NACC cohorts had available harmonized domain-specific cognitive scores (N = 3788 memory, N = 3746 executive, N = 3790 language, and N = 1178 visuospatial). Presence or absence of DLB core features was available for 462 participants across NACC, E-DLB, Pre-D, and ViPD (N = 457 visual hallucinations, N = 375 RBD, N = 364 cognitive fluctuations, N = 335 parkinsonism). Among participants having all DLB core features recorded (56% of the 462), 83.4% displayed two or more.

Biomarker data was available for ADNI and NACC participants. ADNI had CSF Aβ (248 CN, 470 MCI, 189 ADD) and CSF p-tau (343 CN, 551 MCI, 197 ADD), Centiloid (295 CN, 277 MCI, 54 ADD) and tau PET cortical SUVR (268 CN, 158 MCI, 54 ADD). Genetic *APOE* ε4 allele counts were available for some (744 CN, 350 MCI, 934 ADD) ADNI participants. Finally, ADNI and NACC provided neuropathological information which we classified following Wei et al.^54^ into AD (N = 220), LBD (N = 34), and AD+LBD (N = 93). For a detailed breakdown see **Table S2**.

**Table 1:**
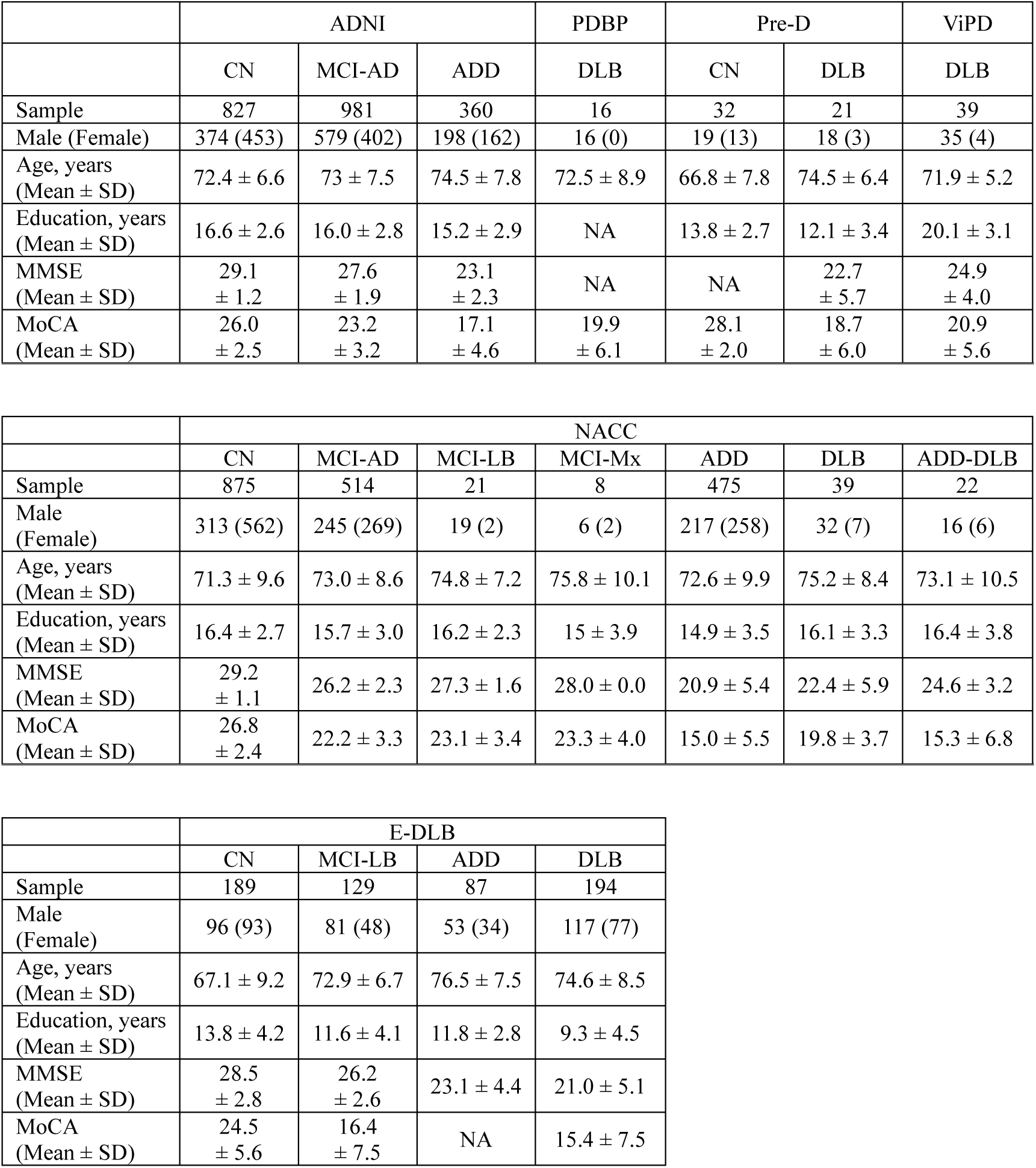
Basic demographic and cognitive information grouped by clinical diagnosis. The MCI group is subdivided into suspected cause. CN = Cognitively Normal, MCI = Mild Cognitive Impairment, MCI-AD: MCI due to probable AD, ADD = Alzheimer’s disease dementia, MCI-LB: MCI due to probable LB, DLB = dementia with Lewy bodies, Mx = Mixed phenotype (meaning ADD and DLB), MMSE = Mini Mental State Examination, MoCA = Montreal Cognitive Assessment, BMK = Biomarker data were used to determine diagnosis.

|  | ADNI |  |  | PDBP | Pre-D |  | ViPD |
| --- | --- | --- | --- | --- | --- | --- | --- |
|  | CN | MCI-AD | ADD | DLB | CN | DLB | DLB |
| Sample | 827 | 981 | 360 | 16 | 32 | 21 | 39 |
| Male (Female) | 374 (453) | 579 (402) | 198 (162) | 16 (0) | 19 (13) | 18 (3) | 35 (4) |
| Age, years<br>(Mean ± SD) | 72.4 ± 6.6 | 73 ± 7.5 | 74.5 ± 7.8 | 72.5 ± 8.9 | 66.8 ± 7.8 | 74.5 ± 6.4 | 71.9 ± 5.2 |
| Education, years<br>(Mean ± SD) | 16.6 ± 2.6 | 16.0 ± 2.8 | 15.2 ± 2.9 | NA | 13.8 ± 2.7 | 12.1 ± 3.4 | 20.1 ± 3.1 |
| MMSE<br>(Mean ± SD) | 29.1<br>± 1.2 | 27.6<br>± 1.9 | 23.1<br>± 2.3 | NA | NA | 22.7<br>± 5.7 | 24.9<br>± 4.0 |
| MoCA<br>(Mean ± SD) | 26.0<br>± 2.5 | 23.2<br>± 3.2 | 17.1<br>± 4.6 | 19.9<br>± 6.1 | 28.1<br>± 2.0 | 18.7<br>± 6.0 | 20.9<br>± 5.6 |

|  | NACC |  |  |  |  |  |  |
| --- | --- | --- | --- | --- | --- | --- | --- |
|  | CN | MCI-AD | MCI-LB | MCI-Mx | ADD | DLB | ADD-DLB |
| Sample | 875 | 514 | 21 | 8 | 475 | 39 | 22 |
| Male<br>(Female) | 313 (562) | 245 (269) | 19 (2) | 6 (2) | 217 (258) | 32 (7) | 16 (6) |
| Age, years<br>(Mean ± SD) | 71.3 ± 9.6 | 73.0 ± 8.6 | 74.8 ± 7.2 | 75.8 ± 10.1 | 72.6 ± 9.9 | 75.2 ± 8.4 | 73.1 ± 10.5 |
| Education, years<br>(Mean ± SD) | 16.4 ± 2.7 | 15.7 ± 3.0 | 16.2 ± 2.3 | 15 ± 3.9 | 14.9 ± 3.5 | 16.1 ± 3.3 | 16.4 ± 3.8 |
| MMSE<br>(Mean ± SD) | 29.2<br>± 1.1 | 26.2 ± 2.3 | 27.3 ± 1.6 | 28.0 ± 0.0 | 20.9 ± 5.4 | 22.4 ± 5.9 | 24.6 ± 3.2 |
| MoCA<br>(Mean ± SD) | 26.8<br>± 2.4 | 22.2 ± 3.3 | 23.1 ± 3.4 | 23.3 ± 4.0 | 15.0 ± 5.5 | 19.8 ± 3.7 | 15.3 ± 6.8 |

|  | E-DLB |  |  |  |
| --- | --- | --- | --- | --- |
|  | CN | MCI-LB | ADD | DLB |
| Sample | 189 | 129 | 87 | 194 |
| Male<br>(Female) | 96 (93) | 81 (48) | 53 (34) | 117 (77) |
| Age, years<br>(Mean ± SD) | 67.1 ± 9.2 | 72.9 ± 6.7 | 76.5 ± 7.5 | 74.6 ± 8.5 |
| Education, years<br>(Mean ± SD) | 13.8 ± 4.2 | 11.6 ± 4.1 | 11.8 ± 2.8 | 9.3 ± 4.5 |
| MMSE<br>(Mean ± SD) | 28.5<br>± 2.8 | 26.2<br>± 2.6 | 23.1 ± 4.4 | 21.0 ± 5.1 |
| MoCA<br>(Mean ± SD) | 24.5<br>± 5.6 | 16.4<br>± 7.5 | NA | 15.4 ± 7.5 |

### The TranSuStaIn Model: atrophy subtypes across ADD and DLB

SuStaIn uncovered two atrophy subtypes in each dementia syndrome, according to the (10-fold) cross validation information criterion.^29^ These subtypes were validated longitudinally, whereby 85.3% of follow-up data is consistently assigned to the same subtype as at baseline (see Supplementary Information: The Null Subtype: Implementation and Implications). The baseline analysis was repeated both with and without batch harmonisation (see Supplementary Information: Data harmonization). Statistical analysis of these four subtypes as described in Methods (see Supplementary Information: SuStaIn subtypes within ADD and DLB) resulted in three of the four being included in our final TranSuStaIn model — one from the DLB model was redundant (see Transdiagnostic Subtyping Framework).

The three TranSuStaIn atrophy subtypes are named for the earliest brain regions to show moderate atrophy (z > 1): Limbic (N = 973), Cortico-Limbic (N = 491), and Cortical (N = 415). A fourth subgroup called Stage Zero (N = 1586) includes individuals assigned to sub-threshold atrophy model stage by SuStaIn. **Figure 2** includes a brain visualisation of the progression patterns, showing left-to-right accumulation of atrophy across regions of interest from subtle (red, z = 0.67) through moderate (magenta, z = 1) and severe atrophy (blue, z = 2). The Limbic subtype shows early moderate atrophy in limbic regions, and only mild early atrophy in cortical regions, while the Cortical subtype atrophy pattern is, broadly speaking, the reverse. The Cortico-Limbic subtype depicts an intermediate pattern of atrophy, where both limbic and cortical atrophy occurs in parallel.

**Figure 1:**
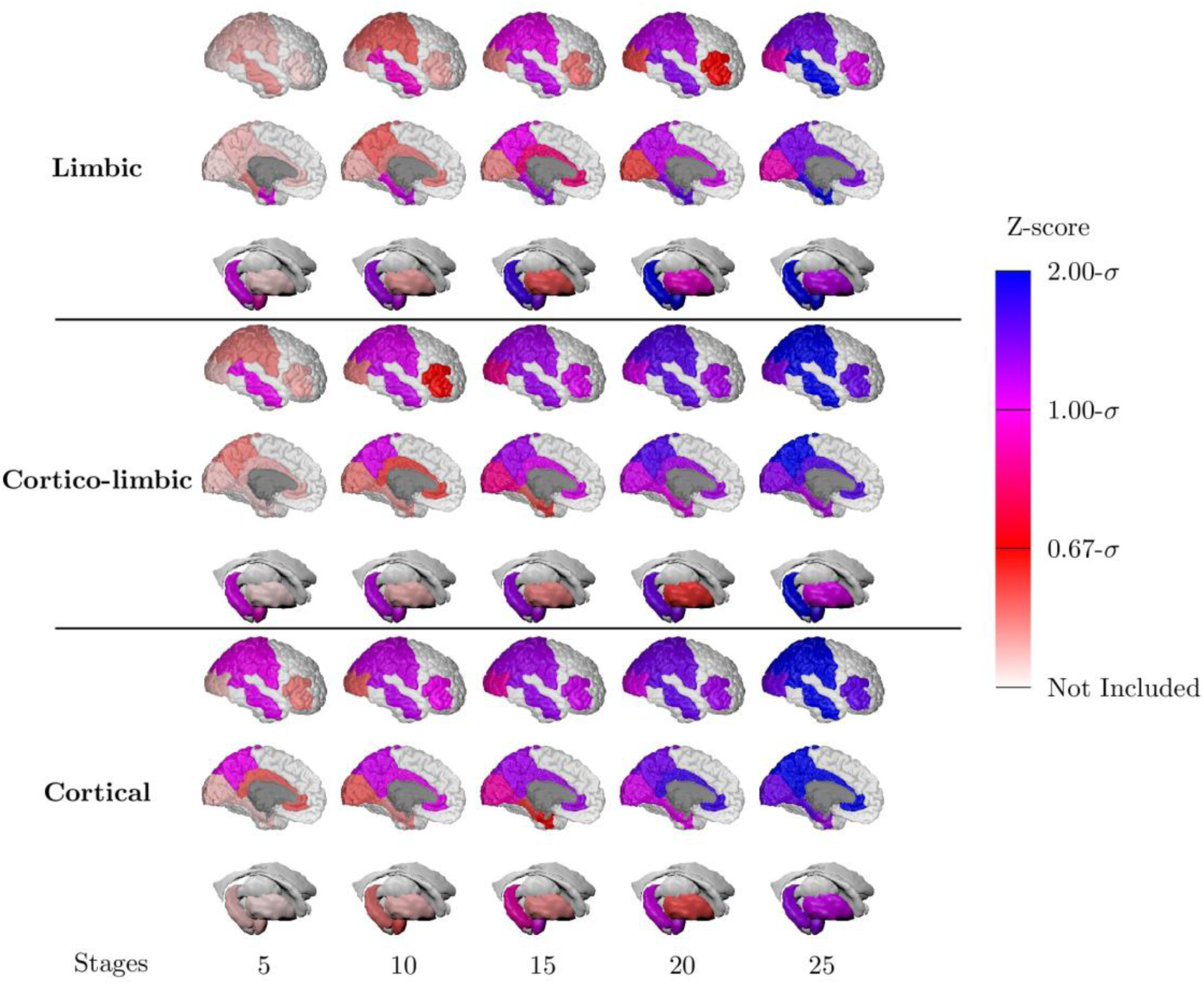
TranSuStaIn uncovered three distinct atrophy patterns. The figure is organized in three rows representing the different subtypes: Limbic, Cortico-Limbic, and Cortical. The brain regions are coloured based on their atrophy z-score level (red: 0.67, magenta: 1, blue: 2), and those regions that were not included are left uncoloured.

### Clinical characteristics of transdiagnostic atrophy subgroups

TranSuStaIn strata were differentially associated with demographic and clinical variables. **Table 2** summarises the statistical comparisons that we present in detail below.

Independent of diagnosis, there was a small but statistically significant difference in sex (female: 42%, 51%, 46%, 54% for Cortical, Cortico-Limbic, Limbic, Stage Zero, respectively: p_FDR_ = 1.70×10^-5^, χ^2^ = 27.7, V = 0.089). Within cognitively impaired individuals only, the small statistical difference remained (34%, 45%, 49%, 44%, respectively: p_FDR_ = 1.20×10^-2^, χ^2^ = 12.8, V = 0.079). Stage Zero were younger (median 71.4) than the atrophy subtypes (median 73.79: p_FDR_ 1.33×10^-16^; MWU = 1.7×10^6^). Within atrophy subtypes, robust linear regression adjusted for model stage (atrophy severity) revealed that the Cortical subtype was the youngest (mean 69.6; CI = [68.0, 71.3] years), followed by Cortico-Limbic (73.8; CI = [72.0, 75.6], p_FDR_ = 3.62×10^-5^), then Limbic (75.7; CI = [74.4, 76.9], p_FDR_ = 8.34×10^-13^). This same analysis revealed subtype-specific age gradients: age of the Cortical subtype increased by 0.2 years per stage (p_FDR_ = 2.07×10^-3^), Cortico-Limbic decreasing by 0.12 years per stage (p_FDR_ = 2.50×10^-2^), and Limbic showing no age gradient.

Clinical diagnosis was statistically and strongly associated with TranSuStaIn atrophy subgroup (p_FDR_ = 2.49×10^-186^, χ^2^ = 887, V = 0.36): ADD is associated with Limbic and Cortico-Limbic subtypes; DLB with Cortical and Cortico-Limbic; and CN with Stage Zero. Cognitive status was associated with model stage (atrophy severity) across all subtypes (Kruskal-Wallis test p_FDR_ < 10^-11^): CN earlier than MCI (Mann-Whitney U test p_FDR_ < .02), and MCI earlier than Dementia (p_FDR_ < 10^-3^).

There were TranSuStaIn subtype associations with cognitive test scores. Using robust linear regression to adjust for model stage and covarying for age and education, general cognition was slightly preserved in the early stages of the Cortical subtype (MMSE = 28) versus the Limbic subtype (MMSE = 27; p_FDR_ = 5.21×10^-3^; values are for age = 73.4 and education = college or higher). Memory was also relatively preserved in the Cortical subtype compared to each of the Limbic and Cortico-Limbic subtypes (memory score difference -0.44, CI = [-0.7, -0.2], p_FDR_ < 10^-4^). No other domain-specific scores showed a statistical association with subtype.

TranSuStaIn subtypes and stages could differentiate only one core feature of DLB: hallucinations. Subtypes did not differ statistically in the relative presence of core features (χ^2^ < 7, p_FDR_ > 0.05), but they did in the presence of hallucinations across early stages in the Cortical (χ^2^ = 11.1, p_FDR_ = .01, V = 0.38) and Limbic (χ^2^ = 11.7, p_FDR_ = .007, V = 0.45) subtypes. Participants in the Cortical subtype at early stages (1–10) had a moderately negative association (residual = -1.4) that reverted at middle stages (11–20, residual = 1.8), while those in the Limbic subtype displayed an opposite pattern (residuals = 1.7 and -1.7, for the early and middle stages respectively).

**Table 2:** Summary statistics of all clinical variables across subtypes. All estimates are displayed as the predicted value and the 99% confidence interval. Comparisons are done across, within, and /or in-between subtypes, and all p-values are FDR corrected. Group analysis shows the result for Kruskal-Wallis or Chi-square tests. The intra-group comparisons of the expected stage for the CN, MCI, and Dementia groups are in the first row within column Kruskal-Wallis tests, and the next two rows Mann-Whitney tests done CN vs MCI and MCI vs Dementia. The intra-group analysis for the slope of the regression analysis assesses the likelihood of being distinct from zero. Pair-wise comparisons are done with-in rows. CI = Cognitively Impaired, CN = Cognitively normal, MCI = Mild Cognitive Impairment, L = Limbic, CL = Cortico-Limbic, C = Cortical.

|  | Limbic | Cortico-Limbic | Cortical | Stage Zero | Group Analysis | Intra-group analysis |  |  | Pair-wise comparisons |  |  |
| --- | --- | --- | --- | --- | --- | --- | --- | --- | --- | --- | --- |
|  |  |  |  |  |  | L | CL | C | L vs CL | CL vs C | C vs L |
| **Biological Sex**<br>(Female/Male) | 449/524 | 249/242 | 175/240 | 861/725 | **p < 0.001** |  |  |  |  |  |  |
| **Biological Sex CI**<br>(Female/Male) | 371/459 | 200/210 | 82/156 | 244/313 | **p < 0.05** |  |  |  |  |  |  |
| **Median Age** | 75.2<br>[51.9, 93.5] | 72.7<br>[49.2, 90.6] | 71.0<br>[52.1, 90.0] | 71.4<br>[48.9, 90.0] | **p < 0.001** |  |  |  |  |  |  |
| **Age and Stage** |  |  |  |  |  |  |  |  |  |  |  |
| Intercept | 75.7<br>[74.4, 76.9] | 73.8<br>[72.0, 75.6] | 69.6<br>[68.0, 71.3] |  |  |  |  |  |  | **p < 0.001** | **p < 0.001** |
| Slope | 0.03<br>[-0.11, 0.06] | -0.12<br>[-0.24, 0.01] | 0.2<br>[0.04, 0.33] |  |  | p > 0.05 | **p < 0.05** | **p < 0.01** |  |  |  |
| **Clinical Diagnosis** |  |  |  |  | **p < 0.001** |  |  |  |  |  |  |
| CN | 143 | 81 | 177 | 1029 |  |  |  |  |  |  |  |
| Probable AD | 761 | 346 | 156 | 488 |  |  |  |  |  |  |  |
| Probable LB | 62 | 56 | 79 | 64 |  |  |  |  |  |  |  |
| Average stage by<br>cognitive status |  |  |  |  |  | **p < 0.001** | **p < 0.001** | **p < 0.001** |  |  |  |
| CN | 5.1<br>[4.0, 6.3] | 7.1<br>[5.1, 9.0] | 5.7<br>[4.8, 6.7] | 5.1<br>[4.0, 6.3] |  | **p < 0.001** | **p < 0.001** | **p < 0.05** |  |  |  |
| MCI | 10.9<br>[9.9, 11.8] | 11.7<br>[10.3, 13.1] | 8.5<br>[6.8, 10.2] | 10.9<br>[9.9, 11.8] |  | **p < 0.001** | **p < 0.001** | **p < 0.001** |  |  |  |
| Dementia | 14.6<br>[13.7, 15.5] | 14.6<br>[13.2, 15.9] | 13.2<br>[11.2, 15.2] | 14.6<br>[13.7, 15.5] |  |  |  |  |  |  |  |
| MMSE and Stage |  |  |  |  |  |  |  |  |  |  |  |
| Intercept | 1.4<br>[0.7, 2.1] | 1.3<br>[0.6, 2.0] | 1.0<br>[0.3, 1.7] |  |  |  |  |  | p > 0.05 | p > 0.05 | **p < 0.01** |
| Slope | 0.024<br>[0.012, 0.035] | 0.028<br>[0.012, 0.044] | 0.033<br>[0.013, 0.053] |  |  | **p < 0.001** | **p < 0.001** | **p < 0.001** | p > 0.05 | p > 0.05 | p > 0.05 |
| Memory and Stage |  |  |  |  |  |  |  |  |  |  |  |
| Intercept | 0.21<br>[-0.31, 0.73] | 0.21<br>[-0.31, 0.73] | 0.65<br>[0.13, 1.17] |  |  |  |  |  | p > 0.05 | **p < 0.001** | **p < 0.001** |
| Slope | -0.023<br>[-0.030, -0.015] | -0.014<br>[-0.025, -0.004] | -0.038<br>[-0.056, -0.020] |  |  | **p < 0.001** | **p < 0.01** | **p < 0.001** | p > 0.05 | **p < 0.01** | p > 0.05 |
| **Executive and Stage** |  |  |  |  |  |  |  |  |  |  |  |
| Intercept | 0.55<br>[-0.10, 1.19] | 0.45<br>[-0.19, 1.09]] | 0.63<br>[-0.01, 1.27] |  |  |  |  |  | p > 0.05 | p > 0.05 | p > 0.05 |
| Slope | -0.017<br>[-0.026, -0.008] | -0.026<br>[-0.039, -0.013] | -0.038<br>[-0.060, -0.016] |  |  | p < 0.001 | p < 0.001 | p < 0.001 | p > 0.05 | p > 0.05 | p = 0.05 |
| **Language and Stage** |  |  |  |  |  |  |  |  |  |  |  |
| Intercept | 1.27<br>[0.72,1.83] | 1.20<br>[0.65, 1.75] | 1.26<br>[0.71, 1.81] |  |  |  |  |  | p > 0.05 | p > 0.05 | p > 0.05 |
| Slope | -0.021<br>[-0.029, -0.013] | -0.020<br>[-0.031, -0.008] | -0.027<br>[-0.046, -0.007] |  |  | p < 0.001 | p < 0.001 | p < 0.01 | p > 0.05 | p > 0.05 | p > 0.05 |
| **Visuospatial and Stage** |  |  |  |  |  |  |  |  |  |  |  |
| Intercept | -0.05<br>[-0.67, 0.58] | 0.06<br>[-0.54, 0.66] | -0.07<br>[-0.71, 0.58] |  |  |  |  |  | p > 0.05 | p > 0.05 | p > 0.05 |
| Slope | -0.001<br>[-0.010, 0.008] | -0.015<br>[-0.026, -0.004] | 0.003<br>[-0.020, 0.026] |  |  | p > 0.05 | p < 0.01 | p > 0.05 | p < 0.05 | p > 0.05 | p > 0.05 |
| **Hallucinations (yes/no)** | 29/28 | 22/29 | 40/37 |  | p > 0.05 | p < 0.01 | p > 0.05 | p < 0.01 |  |  |  |
| Stage (0, 10] | 18/6 | 11/11 | 14/25 |  |  |  |  |  |  |  |  |
| Stage (10, 20] | 5/16 | 5/14 | 17/4 |  |  |  |  |  |  |  |  |
| Stage (20, 25] | 6/6 | 6/4 | 9/8 |  |  |  |  |  |  |  |  |
| **Parkinsonism (yes/no)** | 36/27 | 20/17 | 23/20 |  | p > 0.05 | p = 0.05 | p > 0.05 | p > 0.05 |  |  |  |
| Stage (0, 10] | 8/8 | 6/9 | 15/12 |  |  |  |  |  |  |  |  |
| Stage (10, 20] | 11/5 | 11/4 | 12/8 |  |  |  |  |  |  |  |  |
| Stage (20, 25] | 4/7 | 3/4 | 9/7 |  |  |  |  |  |  |  |  |
| **RBD (yes/no)** | 41/8 | 27/17 | 44/21 |  | p > 0.05 | p > 0.05 | p > 0.05 | p > 0.05 |  |  |  |
| Stage (0, 10] | 19/3 | 9/8 | 24/10 |  |  |  |  |  |  |  |  |
| Stage (10, 20] | 14/5 | 11/7 | 10/6 |  |  |  |  |  |  |  |  |
| Stage (20, 25] | 8/0 | 7/2 | 10/5 |  |  |  |  |  |  |  |  |
| **Cognitive**<br>**Fluctuations (yes/no)** | 34/19 | 25/11 | 45/21 |  | p > 0.05 | p > 0.05 | p > 0.05 | p > 0.05 |  |  |  |
| Stage (0, 10] | 16/5 | 7/7 | 21/10 |  |  |  |  |  |  |  |  |
| Stage (10, 20] | 12/8 | 12/2 | 12/7 |  |  |  |  |  |  |  |  |
| Stage (20, 25] | 6/6 | 6/2 | 12/4 |  |  |  |  |  |  |  |  |

### Genetic, Biological, and Neuropathological characteristics

TranSuStaIn subtypes differed by *APOE* ε4 status, AT(N) status, CSF p-tau/Aβ ratio profiles, and AD neuropathological confirmation (ADNC). Group level analysis shows that the more ADD-like subtypes (Limbic and Cortico-Limbic) had higher prevalence of *APOE* ε4 counts, amyloid and tau positivity, positive CSF ratio increment with stage, and disproportionate assignment of mid-to-high ADNC when compared to the Cortical (more DLB-like) subtype. SAA+ status prevalence was similarly low across subtypes. **Figure 2** summarises the main findings that we present in detail below.

*APOE* ε4 status (0/1/2; available for N = 789) differed across subtypes (see **Figure 2a**; χ^2^ = 28.2, p_FDR_ = 2.04×10^-6^, V = 0.134). The Cortical subtype showed a strong negative association with allele prevalence (residual = -0.88 and -3.61, for 1 and 2 allele count respectively) compared to the Cortico-Limbic (residual = -0.17 and 0.92) and Limbic (residual = 0.67 and 1.50). Conversely, absence of the allele was positively associated with the Cortical subtype (residual = 2.9) and moderately and negatively associated with the Cortico-Limbic and Limbic (residuals = -0.38 and -1.47, respectively) subtypes.

A/T status (in the ATN framework) measured via CSF (see Supplementary Information: PET biomarker profiles in early stages across subtypes for analysis on PET data across subtypes) differed across subtypes (N = 403, χ^2^ = 39.5, p_FDR_ = 3.91×10^-7^, V = 0.22, see **Figure 2b**). The Cortical subtype demonstrated strong positive association with A-T-status (residual = 4.6), while the Cortico-Limbic and Limbic subtypes showed an opposite association – with the Cortico-Limbic subtype showing stronger association (residuals = -2.2 and -0.74). The Cortico-Limbic subtype was positively associated with the A+T+ status (residual = 1.81) while the Limbic subtype was positively associated with A+T-status (residual = 1.12).

TranSuStaIn subtypes and stages showed distinct CSF p-tau/Aβ ratio profiles. Participants in the Cortical subtype had a mean ratio of 0.029 at the early stages (N = 61, CI = [0.011, 0.078]), higher and statistically significant when compared to the Limbic (N = 211, early stage ratio = 0.045, 99% CI = [0.017, 0.123], p_FDR_ = 0.031) and Cortico-Limbic (N = 131, early stage ratio = 0.043, 99% CI = [0.016, 0.118], p_FDR_ = 0.026, see **Figure S9**). These differences in early-stage ratio levels disappear when we control for *APOE* ε4 allele count, suggesting that genetic risk conveyed by *APOE* ε4 explains much of the subtype variance in CSF p-tau/Aβ ratio. Nevertheless, when more data is included – we account for follow-up visits of those subjects whose data from the initial visit was assigned to the Null subtype – the differences between the protein levels in the Cortical and Cortico-Limbic subtypes is no longer explained by the genetic differences alone, suggesting that in the main analysis we lacked power (see Supplementary Information: Extended Analysis: Genetic and CSF including first non-Null visit).

There were no discernible statistical differences in SAA status (N = 223: 58 SAA+, 165 SAA-) across TranSuStaIn subtypes. Using χ^2^ contingency test we found no association between SAA status and phenotype (p_FDR_ = 0.74, χ^2^ = 0.912, V = 0.064), with the prevalence of SAA+ across subtypes being 24.1%, 30.9%, and 24.5% for the Cortical, Cortico-Limbic and Limbic subtypes respectively. These results were reproduced when we included non-Null visits (see Supplementary Information: Extended Analysis: Genetic and CSF including first non-Null visit).

TranSuStaIn subtypes were associated with neuropathological diagnosis (see **Figure 2c**). In the neuropathologically confirmed subset (N = 200) most participants (91%) had ADNC middle to high, which is sufficient to determine that AD could explain the cognitive decline. These participants were disproportionally assigned more to the Limbic subtype (residual = 5.05; χ^2^ = 54.2, p_FDR_ = 1.61×10^-11^, V = 0.386), and less to the Cortical subtype (residual = -5.35).

In summary, genetic risk for AD, CSF A+/T+ status, and neuropathological confirmation of AD are all positively associated with the Cortico-Limbic and Limbic subtypes and negatively associated with the Cortical subtype.

**Figure 2:**
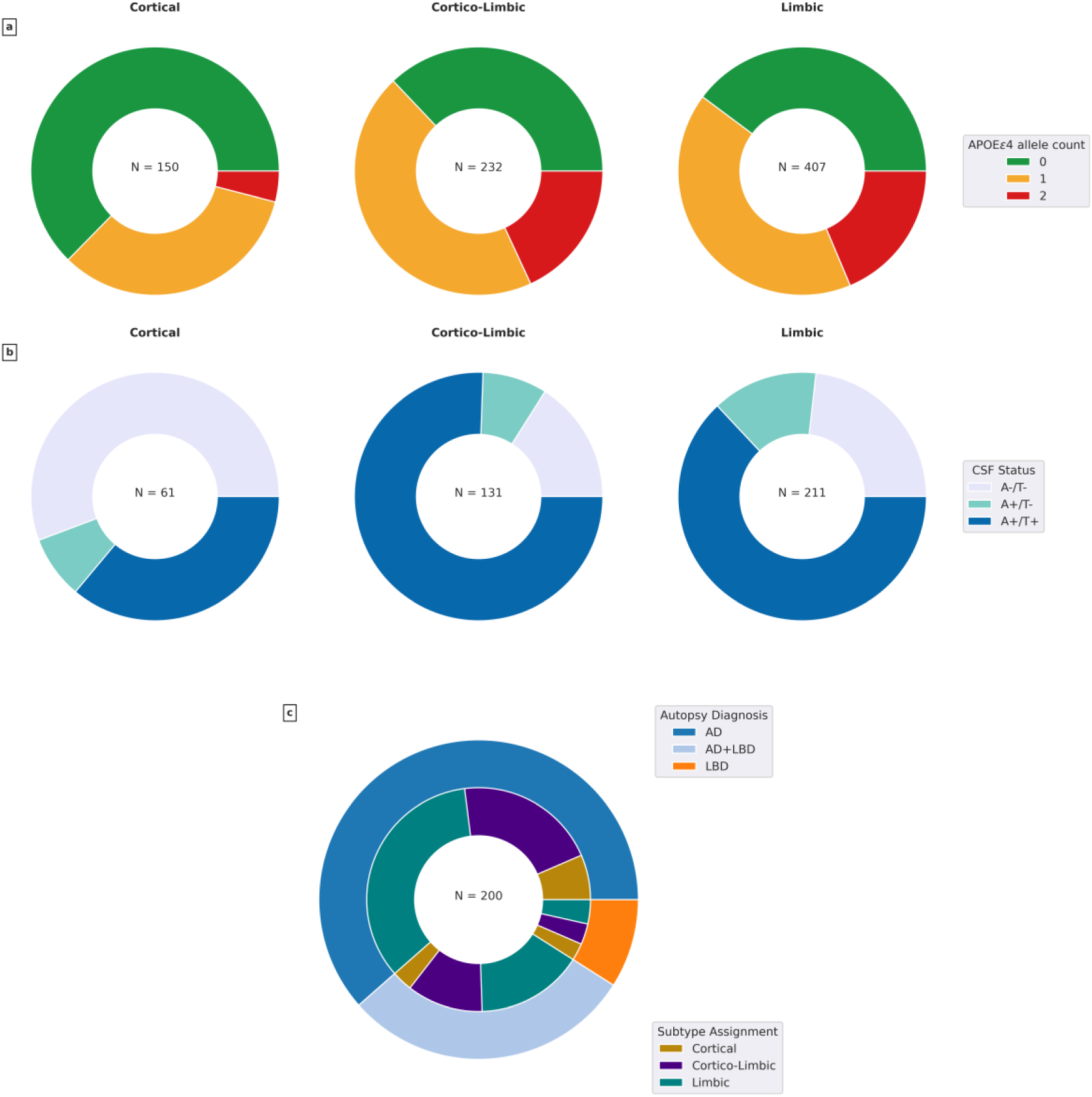
TranSuStaIn model subtype associations with *APOE* ε4 status (panel a), AT(N) status (panel b), and histopathological diagnosis (panel c). **a** Shows the proportion of *APOE* ε4 carriers in each subtype. **b** Shows the proportion positivity across Amyloid and Tau according to the CSF p-tau/Aβ ratio. **c** Shows the subtype assignments for subjects with a histopathological diagnosis of AD, AD+LBD, and LBD. A-/T-= Amyloid and Tau negative, A+/T-= Amyloid positive and Tau negative, A+/T+ = Amyloid and Tau positive, AD = Alzheimer’s disease, LBD = Lewy body disease

## Discussion

We used a commonly available neuroimaging modality (T1w MRI) and augmented a state-of-the-art disease progression modelling algorithm (SuStaIn), to build a transdiagnostic data-driven model of spatiotemporal brain atrophy subtypes across two dementia syndromes: ADD and DLB. Our statistical analysis revealed significant differences (and similarities) across clinical, genetic, biological, and neuropathological features of these atrophy subtypes. Here we discuss implications of our findings in the context of previous work, with a view to assessing clinical utility of our transdiagnostic, data-driven approach.

Our TranSuStaIn model consists of three spatiotemporal brain atrophy signatures/profiles, named by the earliest brain regions to show moderate atrophy: Limbic, Cortico-Limbic, and Cortical. There are qualitative similarities between our transdiagnostic subtypes and previous work on biological subtypes, or biotypes, of ADD.^27^ The “limbic-first” biotype qualitatively aligns with our Limbic subtype, the “typical AD” biotype aligns with our Cortico-Limbic subtype, and the “hippocampal sparing” biotype aligns with our Cortical subtype. While we are not aware of previous work on biotypes of atrophy in DLB/LBD, our Cortical subtype was more “DLB-like”, which is also consistent with previous work on classifying DLB cases from controls using visual ratings.^28^ SuStaIn has been previously applied on *in vivo* MRI-based atrophy features in ADD and DLB,^29,33,55–57^ but not transdiagnostically. Direct comparison with previous work is not straightforward due to the different ROI-based features used but there are similarities, but each study found some combination of Limbic, Cortical, and Subcortical atrophy patterns using MRI volumes and/or cortical thickness features.

Clinically, participants in the Cortical subtype were more likely to have a DLB diagnosis and preserved memory compared to the other subtypes. This was a modest, cross-sectional effect (+1 MMSE point on average) with no statistically significant subtype difference in the rate of general cognitive decline as a function of model stage. Model stage represents cumulative atrophy but is pseudo-longitudinal and so we would require intra-individual longitudinal data to compare with previous longitudinal studies showing divergent rates of cognitive decline between ADD and DLB.^58,59^ Additionally, age and stage interactions in the Cortical subtype showed a positive association, while the Cortico-Limbic subtype demonstrated a negative association. These age and disease severity associations could partially be explained by the relatively low prevalence of early-onset cases in DLB, and relatively high proportion in ADD populations.^60^

Biologically, participants in the Limbic and Cortico-Limbic subtypes were more likely to be *APOE* ε4 positive and have AD-like pathology both *in vivo* (higher CSF p-tau/Aβ ratio) and postmortem (medium to high ADNC), than participants in the Cortical subtype. Higher prevalence of *APOE ε*4 has previously been reported in AD and AD+LBD compared to pure LBD.^61^ Likewise, previous work in neuropathologically confirmed LBD cases who were clinically diagnosed as ADD during life, reported lower tau (higher amyloid) levels.^10,11,13^ CSF markers of α-syn were available for a subset of participants diagnosed with ADD, but we did not find an association with subtype despite the Cortico-Limbic subtype displaying higher rates of SAA+ with atrophy severity. Previous work on ADNI reported differential performance across LBD localization (amygdala-predominant, limbic, or neocortical) that could partially explain our results.^38^ Finally, we found statistical alignment of neuropathological confirmation and subtype, with mid to high ADNC significantly associated with the Cortico-Limbic and Limbic subtypes but not the Cortical subtype.

Our results are consistent with the prevailing idea that AD (and ADD) atrophy profiles seem to be “more limbic”, DLB (and LBD) atrophy profiles seem to be “more cortical”, and mixed pathology is likely to result in a hybrid Cortico-Limbic profile of spatiotemporal atrophy with a more aggressive progression. In total, this amounts to significant overlap of both syndrome and pathology (both *in vivo* and postmortem) with the data-driven atrophy subtypes in TranSuStaIn. These results support the hypothesis that the spatial signature of brain atrophy in dementias is driven by underlying biological processes/diseases, concordant with the co-pathology profile. Our results also support the notion that our TranSuStaIn model could have stratification utility, although this conclusion would be made stronger by future validation studies involving more complete biomarker profiling, i.e., without the confound of missing data (see discussion of limitations below).

The primary limitation of our study is missing data, especially across biomarkers used to assess disease pathology *in vivo*. This limits our ability to draw strong conclusions about the balance of pathologies underpinning our transdiagnostic atrophy subtypes. Ideally this would be done *within subtype*: on complete biomarker data, in a sample statistically balanced by diagnosis to facilitate comparison of pathologies driving the atrophy signature. Here, missing biomarker data limited us to comparing atrophy subtypes *within pathology* (and within syndrome), which is informative but suboptimal. This is an unavoidable limitation in secondary data analyses such as ours: each data-generating study was enriched for a single dementia syndrome, in some cases supported by biomarker confirmation of a single underlying pathology. The ultimate validation of transdiagnostic approaches like ours requires future prospective studies having comprehensive biomarker panels, neuroimaging, and neuropsychological assessments. Ideally, we might include such measurements directly into our model as input features.

There are other limitations that motivate future work. First, atrophy measured by T1w MRI may lack pathological specificity, although the subtype-pathology associations we found suggests that accounting for the spatial signature of atrophy (as SuStaIn does) partially addresses this limitation. Second, spatial resolution of atrophy subtyping could be improved beyond the ROI-level used here, which was necessitated by computational limitations of the pySuStaIn software package.^62^ This may be achievable via a recent methodological advancement in event-based modelling.^63^ Third, our transdiagnostic approach might be attempted using emerging innovations to SuStaIn like AddiPath that attempt to directly model the spatial signature of co-pathology via an additional imaging modality.^64^

It is increasingly apparent that dementia syndromes are underpinned by complex co-pathologies, necessitating a shift towards more holistic approaches for understanding the neurodegenerative diseases involved.^20,65^ Unfortunately, syndrome-specific studies tend to collect only partially overlapping sets of measurements, which makes it almost impossible to investigate co-pathologies transdiagnostically. Our study is the first to attempt this using SuStaIn exclusively on atrophy-based signal. Importantly, our results on the incomplete biomarker data suggest that MRI based atrophy patterns correlate with distinct biological profiles, possibly enabling *in vivo* differentiation of neurodegenerative disease with TranSuStaIn. To confirm this, future studies should incorporate other neurodegenerative diseases, e.g., frontotemporal lobar degeneration, along with biomarker data. We are optimistic that approaches like TranSuStaIn might soon provide information on underlying disease/copathology in the absence of fluid or PET biomarkers, for playing a data-driven role in clinical care and treatment decisions.

## Author contributions (ICMJE)

GCL: conception, data, analysis, coding, collaboration and discussion, writing – original draft, writing – review and editing, figures.

AK: collaboration and discussion, writing – review and editing.

ALY: collaboration and discussion, writing – review and editing.

NZ: data, writing – review and editing.

TS: data, writing – review and editing.

AH: data, writing – review and editing.

NCP: data, writing – review and editing.

MCG: data, writing – review and editing.

J-PT: data, writing – review and editing.

MF: data, writing – review and editing.

AT: data, writing – review and editing.

PD: data, writing – review and editing.

JO: data, writing – review and editing.

SJGL: data, writing – review and editing.

DA: data, writing – review and editing.

AB: data, writing – review and editing.

APi: data, writing – review and editing.

APa: data, writing – review and editing.

EL: data, writing – review and editing.

2^nd^A: data, writing – review and editing.

KS: data, writing – review and editing.

IR: data, writing – review and editing.

ATI: data, writing – review and editing.

BS: data, writing – review and editing.

CC: data, writing – review and editing.

CL: data, writing – review and editing.

RB: data, writing – review and editing.

ID: data, writing – review and editing.

RSW: data, writing – review and editing.

ZW: data, writing – review and editing.

DAa: data, writing – review and editing.

EW: data, writing – review and editing.

DF: data, writing – review and editing.

EM: conception, data, funding acquisition, project administration, collaboration and discussion, writing – review and editing.

NPO: conception, data, funding acquisition, project administration, collaboration and discussion, writing – original draft, writing – review and editing.

All authors approve of the submitted version of the manuscript.

## Acknowledgements

The authors acknowledge members of the UCL POND group (http://pond.cs.ucl.ac.uk) for their valuable input and feedback received during group discussions.

Data collection and sharing for the Alzheimer’s Disease Neuroimaging Initiative (ADNI) is funded by the National Institute on Aging (National Institutes of Health Grant U19 AG024904). The grantee organization is the Northern California Institute for Research and Education. In the past, ADNI has also received funding from the National Institute of Biomedical Imaging and Bioengineering, the Canadian Institutes of Health Research, and private sector contributions through the Foundation for the National Institutes of Health (FNIH) including generous contributions from the following: AbbVie, Alzheimer’s Association; Alzheimer’s Drug Discovery Foundation; Araclon Biotech; BioClinica, Inc.; Biogen; Bristol-Myers Squibb Company; CereSpir, Inc.; Cogstate; Eisai Inc.; Elan Pharmaceuticals, Inc.; Eli Lilly and Company; EuroImmun; F. Hoffmann-La Roche Ltd and its affiliated company Genentech, Inc.; Fujirebio; GE Healthcare; IXICO Ltd.; Janssen Alzheimer Immunotherapy Research & Development, LLC.; Johnson & Johnson Pharmaceutical Research &Development LLC.; Lumosity; Lundbeck; Merck & Co., Inc.; Meso Scale Diagnostics, LLC.; NeuroRx Research; Neurotrack Technologies; Novartis Pharmaceuticals Corporation; Pfizer Inc.; Piramal Imaging; Servier; Takeda Pharmaceutical Company; and Transition Therapeutics.

The NACC database is funded by NIA/NIH Grant U24 AG072122. NACC data are contributed by the NIA-funded ADRCs: P30 AG062429 (PI James Brewer, MD, PhD), P30 AG066468 (PI Oscar Lopez, MD), P30 AG062421 (PI Teresa Gomez-Isla, MD), P30 AG066509 (PI Thomas Grabowski, MD), P30 AG066514 (PI Mary Sano, PhD), P30 AG066530 (PI Helena Chui, MD, Arthur Toga, PhD), P30 AG066507 (PI Marilyn Albert, PhD), P30 AG066444 (PI David Holtzman, MD), P30 AG066518 (PIs Lisa Silbert, MD, Kevin Duff, PhD), P30 AG066512 (PI Thomas Wisniewski, MD), P30 AG066462 (PI Scott Small, MD), P30 AG072979 (PI David Wolk, MD), P30 AG072972 (PIs Charles DeCarli, MD, Rachel Whitmer, PhD), P30 AG072976 (PI Andrew Saykin, PsyD), P30 AG072975 (PI Julie Schneider, MD, MS), P30 AG072978 (PI Ann McKee, MD), P30 AG072977 (PI Robert Vassar, PhD), P30 AG066519 (PI Joshua Grill, PhD), P30 AG062677 (PIs Brad Boeve, MD, Ronald Petersen, MD, PhD), P30 AG079280 (PI Jessica Langbaum, PhD), P30 AG062422 (PI Gil Rabinovici, MD), P30 AG066511 (PI Allan Levey, MD, PhD), P30 AG072946 (PI Linda Van Eldik, PhD), P30 AG062715 (PI Sanjay Asthana, MD, FRCP), P30 AG072973 (PI Russell Swerdlow, MD), P30 AG066506 (PIs Glenn Smith, PhD, ABPP, David Lowenstein, PhD, Ranjan Duara, MD), P30 AG066508 (PIs Stephen Strittmatter, MD, PhD, Christopher Van Dyck, MD), P30 AG066515 (PI Victor Henderson, MD, MS), P30 AG072947 (PI Suzanne Craft, PhD), P30 AG072931 (PI Henry Paulson, MD, PhD), P30 AG066546 (PIs Sudha Seshadri, MD, Gladys Maestre, MD, PhD), P30 AG086401 (PI Erik Roberson, MD, PhD), P30 AG086404 (PI Gary Rosenberg, MD), P30 AG086403 (PI Angela Jefferson, PhD), P30 AG072958 (PIs Heather Whitson, MD, Gwenn Garden, MD, PhD), P30 AG072959 (PI Jagan Pillai, MD, PhD), P30 AG092752 (Ihab Hajjar, MD, MS).

The NACC database is funded by NIA/NIH Grant U24 AG072122. SCAN is a multi-institutional project that was funded as a U24 grant (AG067418) by the National Institute on Aging in May 2020. Data collected by SCAN and shared by NACC are contributed by the NIA-funded ADRCs as follows:

Arizona Alzheimer’s Center - P30 AG072980 (PI: Eric Reiman, MD); R01 AG069453 (PI: Eric Reiman (contact), MD); P30 AG019610 (PI: Eric Reiman, MD); and the State of Arizona which provided additional funding supporting our center; Boston University - P30 AG013846 (PI Neil Kowall MD); Cleveland ADRC - P30 AG062428 (James Leverenz, MD); Cleveland Clinic, Las Vegas – P20AG068053; Columbia - P50 AG008702 (PI Scott Small MD); Duke/UNC ADRC – P30 AG072958; Emory University - P30AG066511 (PI Levey Allan, MD, PhD); Indiana University - R01 AG19771 (PI Andrew Saykin, PsyD); P30 AG10133 (PI Andrew Saykin, PsyD); P30 AG072976 (PI Andrew Saykin, PsyD); R01 AG061788 (PI Shannon Risacher, PhD); R01 AG053993 (PI Yu-Chien Wu, MD, PhD); U01 AG057195 (PI Liana Apostolova, MD); U19 AG063911 (PI Bradley Boeve, MD); and the Indiana University Department of Radiology and Imaging Sciences; Johns Hopkins - P30 AG066507 (PI Marilyn Albert, Phd.); Mayo Clinic - P50 AG016574 (PI Ronald Petersen MD PhD); Mount Sinai - P30 AG066514 (PI Mary Sano, PhD); R01 AG054110 (PI Trey Hedden, PhD); R01 AG053509 (PI Trey Hedden, PhD); New York University - P30AG066512-01S2 (PI Thomas Wisniewski, MD); R01AG056031 (PI Ricardo Osorio, MD); R01AG056531 (PIs Ricardo Osorio, MD; Girardin Jean-Louis, PhD); Northwestern University - P30 AG013854 (PI Robert Vassar PhD); R01 AG045571 (PI Emily Rogalski, PhD); R56 AG045571, (PI Emily Rogalski, PhD); R01 AG067781, (PI Emily Rogalski, PhD); U19 AG073153, (PI Emily Rogalski, PhD); R01 DC008552, (M.-Marsel Mesulam, MD); R01 AG077444, (PIs M.-Marsel Mesulam, MD, Emily Rogalski, PhD); R01 NS075075 (PI Emily Rogalski, PhD); R01 AG056258 (PI Emily Rogalski, PhD); Oregon Health and Science University - P30 AG066518 (PIs Lisa Silbert, MD, Kevin Duff, PhD); R56 AG074321 (PI Jeffrey Kaye, MD); Rush University - P30 AG010161 (PI David Bennett MD); Stanford – P30AG066515; P50 AG047366 (PI Victor Henderson MD MS); University of Alabama, Birmingham – P20; University of California, Davis - P30 AG10129 (PI Charles DeCarli, MD); P30 AG072972 (PI Charles DeCarli, MD); University of California, Irvine - P50 AG016573 (PI Frank LaFerla PhD); University of California, San Diego - P30AG062429 (PI James Brewer, MD, PhD); University of California, San Francisco - P30 AG062422 (Rabinovici, Gil D., MD); University of Kansas - P30 AG035982 (Russell Swerdlow, MD); University of Kentucky - P30 AG028283-15S1 (PIs Linda Van Eldik, PhD and Brian Gold, PhD); University of Michigan ADRC - P30AG053760 (PI Henry Paulson, MD, PhD) P30AG072931 (PI Henry Paulson, MD, PhD) Cure Alzheimer’s Fund 200775 - (PI Henry Paulson, MD, PhD) U19 NS120384 (PI Charles DeCarli, MD, University of Michigan Site PI Henry Paulson, MD, PhD) R01 AG068338 (MPI Bruno Giordani, PhD, Carol Persad, PhD, Yi Murphey, PhD) S10OD026738-01 (PI Douglas Noll, PhD) R01 AG058724 (PI Benjamin Hampstead, PhD) R35 AG072262 (PI Benjamin Hampstead, PhD) W81XWH2110743 (PI Benjamin Hampstead, PhD) R01 AG073235 (PI Nancy Chiaravalloti, University of Michigan Site PI Benjamin Hampstead, PhD) 1I01RX001534 (PI Benjamin Hampstead, PhD) IRX001381 (PI Benjamin Hampstead, PhD); University of New Mexico - P20 AG068077 (Gary Rosenberg, MD); University of Pennsylvania - State of PA project 2019NF4100087335 (PI David Wolk, MD); Rooney Family Research Fund (PI David Wolk, MD); R01 AG055005 (PI David Wolk, MD); University of Pittsburgh - P50 AG005133 (PI Oscar Lopez MD); University of Southern California - P50 AG005142 (PI Helena Chui MD); University of Washington - P50 AG005136 (PI Thomas Grabowski MD); University of Wisconsin - P50 AG033514 (PI Sanjay Asthana MD FRCP); Vanderbilt University – P20 AG068082; Wake Forest - P30AG072947 (PI Suzanne Craft, PhD); Washington University, St. Louis - P01 AG03991 (PI John Morris MD); P01 AG026276 (PI John Morris MD); P20 MH071616 (PI Dan Marcus); P30 AG066444 (PI John Morris MD); P30 NS098577 (PI Dan Marcus); R01 AG021910 (PI Randy Buckner); R01 AG043434 (PI Catherine Roe); R01 EB009352 (PI Dan Marcus); UL1 TR000448 (PI Brad Evanoff); U24 RR021382 (PI Bruce Rosen); Avid Radiopharmaceuticals / Eli Lilly; Yale - P50 AG047270 (PI Stephen Strittmatter MD PhD); R01AG052560 (MPI: Christopher van Dyck, MD; Richard Carson, PhD); R01AG062276 (PI: Christopher van Dyck, MD); 1Florida - P30AG066506-03 (PI Glenn Smith, PhD); P50 AG047266 (PI Todd Golde MD PhD)

Data and biospecimens used in preparation of this manuscript were obtained from the Parkinson’s Disease Biomarkers Program (PDBP) Consortium, supported by the National Institute of Neurological Disorders and Stroke at the National Institutes of Health. Investigators include: Roger Albin, Roy Alcalay, Alberto Ascherio, Thomas Beach, Sarah Berman, Bradley Boeve, F. DuBois Bowman, Shu Chen, Alice Chen-Plotkin, William Dauer, Ted Dawson, Paula Desplats, Richard Dewey, Ray Dorsey, Jori Fleisher, Kirk Frey, Douglas Galasko, James Galvin, Dwight German, Steven Gunzler, Lawrence Honig, Xuemei Huang, David Irwin, Un Kang, Kejal Kantarci, Anumantha Kanthasamy, Daniel Kaufer, Horacio Kaufmann, Qingzhong Kong, James Leverenz, Allan Levey, Carol Lippa, Irene Litvan, Oscar Lopez, Jian Ma, Richard Mailman, Lara Mangravite, Karen Marder, Kelly Mills, Nandakumar Narayanan, Laurie Orzelius, Vladislav Petyuk, Judith Potashkin, Liana Rosenthal, Rachel Saunders-Pullman, Clemens Scherzer, Michael Schwarzschild, Nicholas Seyfried, Tanya Simuni, Andrew Singleton, David Standaert, Debby Tsuang, David Vaillancourt, Jerrold Vitek, David Walt, Andrew West, Cyrus Zabetian, and Jing Zhang. The PDBP Investigators have not participated in reviewing the data analysis or content of the manuscript

## Funding acknowledgements

This work was made possible by an Ignition grant from the University of Sydney and University College London (Global Engagement Fund), and a travel grant from Alzheimer’s Research UK.

GCL and NPO acknowledge funding from the UKRI Medical Research Council (MR/S03546X/1, MR/X024288/1, MR/T046422/1).

EM was supported by a National Health and Medical Research Council (NHMRC) grant (2008565).

RLR is supported by an NHRMC Emerging Leadership Grant (GNT2010064). JPT is supported by the NIHR Newcastle Biomedical Research Centre.

RSW is supported by a Wellcome Career Development Award (#225263/Z/22/Z), Parkinson’s UK, the Lewy Body Society, Michael J Fox Foundation, and by the National Institute for Health and Care Research University College London Hospitals Biomedical Research Centre.

CL was supported by the Medical Research Council (MR/R006504/1), Parkinson’s UK, Hilary-Galen Weston Foundation and Michael J Fox Foundation.

SJGL was supported by National Health and Medical Research Council fellowship grant (#1195830) and the NHMRC/EU Joint Programme on Neurodegenerative Disease Research (#2014513).

DA work was supported by research grants from Institute of Health Carlos III (ISCIII), Spain, PI18/00435, PI22/00611, INT19/00016, INT23/00048 to DA, and by the Department of Health Generalitat de Catalunya PERIS program SLT006/17/125 to DA.

AB acknowledges support from Instituto de Salud Carlos III and co-funded by the European Union through the Miguel Servet grant (CP20/00038) and Fondo de Investigaciones Sanitario (PI22/00307), the Alzheimer’s Association (AARG-22-923680), and the Ajuntament de Barcelona, in collaboration with Fundació La Caixa (23S06157-001).

APi has been partially supported by I) #NEXTGENERATIONEU (NGEU) funded by the Ministry of University and Research (MUR), National Recovery and Resilience Plan (NRRP), project MNESYS (PE0000006) – a multiscale integrated approach to the study of the nervous system in health and disease (DN. 1553 11.10.2022) – subproject DIGI-BRAIN II) the European Union -Next Generation EU - NRRsP M6C2 - Investment

2.1 Enhancement and strengthening of biomedical research in the NHS-PNRR – PNRR PNRR-Health PNRR-MAD-2022-12376110; APi has been supported by grants of Airalzh Foundation AGYR2021 Life-Bio Grant, The LIMPE-DISMOV Foundation Segala Grant 2021, the Italian Ministry of University and Research PRIN COCOON (2017MYJ5TH) and PRIN 2021 RePlast (20202THZAW), PRIN 2022PNJS5Z and PRIn PNRR (P20224ZHM9), DIGI-BRAIN the H2020 IMI IDEA-FAST (ID853981),

Italian Ministry of Health, Grant/Award Number: RF-2018-12366209, PNRR-Health PNRR-MAD-2022-12376110 and PNRR-MCNT2-2023-12378387, The MJFF Foundation Grant 022343.

APa has been partially supported by I) #NEXTGENERATIONEU (NGEU) funded by the Ministry of University and Research (MUR), National Recovery and Resilience

Plan (NRRP), project MNESYS (PE0000006) – a multiscale integrated approach to the study of the nervous system in health and disease (DN. 1553 11.10.2022) – subproject DIGI-BRAIN II) the European Union -Next Generation EU - NRRP M6C2 – Investment 2.1 Enhancement and strengthening of biomedical research in the NHS-PNRR – PNRR PNRR-Health PNRR-MAD-2022-12376110; APa has been supported by grants of the Italian Ministry of University and Research PRIN COCOON (2017MYJ5TH) and PRIN 2021 RePlast (20202THZAW), Prin 2022 EGADi (P2022TKN8C) the H2020 IMI

IDEA-FAST (ID853981), DIGI-BRAIN Italian Ministry of Health, Grant/Award Number: RF-2018-12366209, RF-2019-12369272 and PNRR-Health PNRR-MAD-2022-12376110.

EW was supported the Swedish Research Council (VR) No. 2016-02282, 2021-01861, 2025-02405; the Center for Innovative Medicine (CIMED) No. FoUI-954459, FoUI-975174; the regional agreement on medical training and clinical research (ALF) between Stockholm County Council and Karolinska Institutet No. FoUI-952838, FoUI-954893; The Swedish Brain Foundation (Hjärnfonden) No. FO2022-0084, FO2024-0239; The Swedish Alzheimer’s Foundation (Alzheimerfonden) No. AF-967495, AF-980387; The Swedish Parkinson’s foundation (Parkinsonfonden) No. 1647/25, 1557/24, 1521/23; EU Innovative Health Initiative Joint Undertaking (IHI JU) AD-RIDDLE and ACCESS AD; King Gustaf V:s and Queen Victorias Foundation; Olle Engkvists Foundation (Olle Engkvists Stiftelse) No. 186-0660, 224-0069.

Annegret Habich was supported by funding from StratNeuro, Demensfonden, Gun and Bertil Stohnes Stiftelsen (2024-029), Stiftelsen för Gamla Tjänarinnor (2023-016), and various grants from Karolinska Institutet (2024-02083).

DF receives funding from the Swedish Research Council (Vetenskapsrådet, grant 2022-00916 and 2025-02984), the Center for Innovative Medicine (CIMED, grants 20200505 and FoUI-988826), the regional agreement on medical training and clinical research of Stockholm Region (ALF Medicine, grants FoUI-962240 and FoUI-987534), the Swedish Brain Foundation (Hjärnfonden FO2023-0261, FO2022-0175, FO2021-0131), the Swedish Alzheimer Foundation (Alzheimerfonden AF-968032, AF-980580, AF-994058, AF-1010553), the Swedish Dementia Foundation (Demensfonden), the Gamla Tjänarinnor Foundation, the Gun och Bertil Stohnes Foundation, the Åke Wiberg Foundation, StratNeuro, Parkinsonfonden, Funding for Research from Karolinska Institutet, Neurofonden, and the Foundation for Geriatric Diseases at Karolinska Institutet, contributions from private bequests and academic agreements with industry. Zuzana Walker received funding from ARUK and Lewy Body Society.

TS was supported by Stiftelsen för Gamla Tjänarinnor (2024-213) and Foundation for Geriatric Diseases at Karolinska Institutet (FS-2025:0013).

JO is supported by the NIHR Cambridge Biomedical Research Centre and the Medical Research Council funded Dementias Platform UK.

AWL receives funding from the Hersenstichting, ZONMW, MJFF and Gieske Strijbis Fonds

MF is supported by the NIHR Newcastle Biomedical Research CentreIrena Rektorova received funding from the project nr. LX22NPO5107 (MEYS): Financed by European Union – Next Generation EU; from the MEYS (EU Joint Programme Neurodegenerative Disease) 9F25001 TRACE-PD: Tracking the mechanisms of disease progression and functional compensation in the early phase of Parkinson’s disease; and from the MEYS (EU Joint Program Neurodegenerative Disease) 8F22005 TACKL-PRED: TACKLing the Challenges of PREsymptomatic Sporadic Dementia.

PCD is supported by the Medical Research Council [grant number MR/W000229/1] and the NIHR Newcastle Biomedical Research Centre.

AK was supported by The Australian Rotary Health/Rotary Club of Belconnen 50th Anniversary PhD Scholarship (SC4968)

This research was funded in whole, or in part by the Wellcome Trust [227341/Z/23/Z]. For the purpose of open access, the author has applied a CC BY public copyright licence to any Author Accepted Manuscript version arising from this submission.

## Disclosures

NPO is a paid consultant for Queen Square Analytics Limited (UK) on unrelated projects. EM has received speaking and writing honoraria for the International Parkinson and Movement Disorders Society, Somnomed, and CSL Seqirus. RSW has received speaking and writing honoraria from GE Healthcare, Bial, Omnix Pharma, and Britannia; and consultancy fees from Therakind and Accenture. DA participated in advisory boards from Fujirebio-Europe, Roche Diagnostics, Grifols S.A. and Lilly, and received speaker honoraria from Fujirebio-Europe, Roche Diagnostics, Nutricia, Krka Farmacéutica S.L., Zambon S.A.U., Neuraxpharm, Alter Medica, Lilly and Esteve Pharmaceuticals S.A. DA declares a filed patent application (WO2019175379 Markers of synaptopathy in neurodegenerative disease). APi received consultancy/speaker fees from Abbvie, Angelini, Bial, Eli Lilly, Lundbeck, Roche and Zambon pharmaceuticals. He acts as consultant as part of advisory Board of Angelini Pharma and BIAL pharmaceutics. APa received personal compensation as a consultant/scientific advisory board member for Biogen, Eisai Eli Lilly, General Healthcare (GE), Lundbeck, Nestlè, Roche. DF consults for BioArctic and has received honoraria from Esteve Pharmaceuticals S.A.

J-PT has received speaking honoraria from GE Healthcare and acted as a consultant for CervoMed and Eisai. ZW acted as a consultant for GE Healthcare.

## Data Availability Statement

Data for this study was sourced from multiple cohorts. Three of them — ADNI, NACC and PDBP — are publicly available via their respective websites. The E-DLB consortium makes data available upon reasonable request. The python implementation of SuStaIn is publicly available in https://github.com/ucl-pond/pySuStaIn.

## Consent Statement

All data were collected with appropriate informed consent under ethically approved clinical studies. Our secondary analysis of these data was approved by the UCL Research Ethics Committee under application 8019/005 and the UCL Department of Computer Science Research Ethics Committee under application UCL-CSREC-209-B.

## Supplementary Information

### Diagnostic criteria

Diagnostic criteria for each dementia syndrome varied slightly by cohort. ADNI diagnostic criteria through all their phases up to ADNI4 is based on clinical assessments following NINCDS-ADRDA criteria and the clinicians judgement to discard any participants where there is suspicion of any other neurodegenerative disease other than ADD.^66^ NACC clinical data adheres to the Uniform Data Set (UDS)^67^ used by the Alzheimer’s Disease Research Centers (ADRC), and the diagnostic criteria can include biomarker information. PDBP recruitment of clinically diagnosed DLB participants (described elsewhere)^68^ followed the 2017 Dementia with Lewy Bodies Consortium Criteria.^1^ E-DLB diagnostic criteria (described elsewhere)^69^ was made according to the 2005 International Consensus Criteria for probable DLB and the ICD 10 for probable ADD based on all available data.^70^ ViPD participants were diagnosed as having probable DLB if they satisfied the 2017 Dementia with Lewy Bodies Consortium Criteria^1^, and reasons for exclusion included a history of traumatic brain injury, or major co-morbid psychiatric or confounding neurological disorders. Pre-D participants were recruited from the Parkinson’s Disease Research Clinic, University of Sydney and assessed by specialist neurologists and diagnosed according to established consensus criteria for Parkinson’s disease^71^ and DLB.^1^ To ensure sufficient scan quality, we excluded anatomical T1w MRI brain scans from ADNI, NACC, PDBP, ViPD, Pre-D cohorts if acquisition type was not 3D or the spatial resolution was not 1.5 mm or better (smaller) in each direction – E-DLB cohort has a post-processing quality control (see ‘Imaging Derived Phenotypes’). Due to the multicohort nature of the study and considering the varying properties of the scanners (field strength, manufacturer) and the acquisition protocols (see **Table S**), we performed our analyses both with and without data harmonization (see ‘Data harmonization’).

### SuStaIn subtypes within ADD and DLB

The SuStaIn algorithm was initially applied to each clinical cohort separately (**Figure S1**). The Positional Variance Diagram visualization in **Figure S1A** depicts color-coded spatiotemporal atrophy (left-right): red (z=0.67), magenta (z=1), and blue (z=2). Based on CVIC values (**Figure S1B**), we identified two distinct subtypes within each population, characterized by their earliest-affected brain regions: Cortical (DLB), Limbic (in both AD and DLB), and Cortico-Limbic (AD). The Cortical subtype progression shown in **Figure S1A**(i) is characterized by widespread cortical involvement (z-score ≥ 1, magenta), with delayed hippocampal, amygdala, entorhinal, and parahippocampal atrophy. The Limbic subtypes in clinical DLB (**Figure S1A**[ii]) and clinical AD (**Figure S1A[**iii]) demonstrated similar progression patterns to each other, showing early moderate atrophy (z-score ≥ 1, magenta) in three key regions: hippocampus, amygdala, and entorhinal cortex. The cortical regions only progressed to moderate atrophy by the middle stages of disease progression. Notably, the DLB-enriched variant in **Figure S1A**(ii) showed earlier involvement of cortical regions (middle temporal and parietal) when compared to the AD-enriched variant. The Cortico-limbic subtype (**Figure S1A**[iv]) begins with atrophy in both cortical (medial temporal and parietal lobe) and limbic (hippocampus, amygdala) regions.

Quantitative comparison using Hellinger distance demonstrated that the Cortical subtype and both Limbic subtypes were the most divergent from each other (0.815 and 0.674 for the comparisons of the Cortical subtype with the AD and DLB limbic variants, respectively). We observed varying degrees of overlap between the other subtypes, with moderate similarity between the Cortico-Limbic (AD) and both DLB subtypes (0.534 and 0.563 for the comparisons of the Cortico-Limbic with the Cortical and Limbic subtypes, respectively). The greatest similarity was observed between the two Limbic subtypes (0.399). We also explored the relationship between the progression patterns in terms of how well the likelihood to one subtype predicts the likelihood to another subtype. The R2 metric was highest for the two limbic variants (0.82 and 0.76, for the AD variant likelihood predicting the DLB variant likelihood and vice versa) and lowest for the Limbic AD and the Cortical subtypes (-0.39 and -1.45). In between these values lay the rest of the comparisons: the Cortico-Limbic and Cortical (0.45 and 0.15), the Cortico-Limbic and Limbic AD (-0.15 and -0.01), the Cortico-Limbic and Limbic DLB (0.26, 0.16), and the Limbic DLB and Cortical (-0.23 and -0.67).

**Figure S1:**
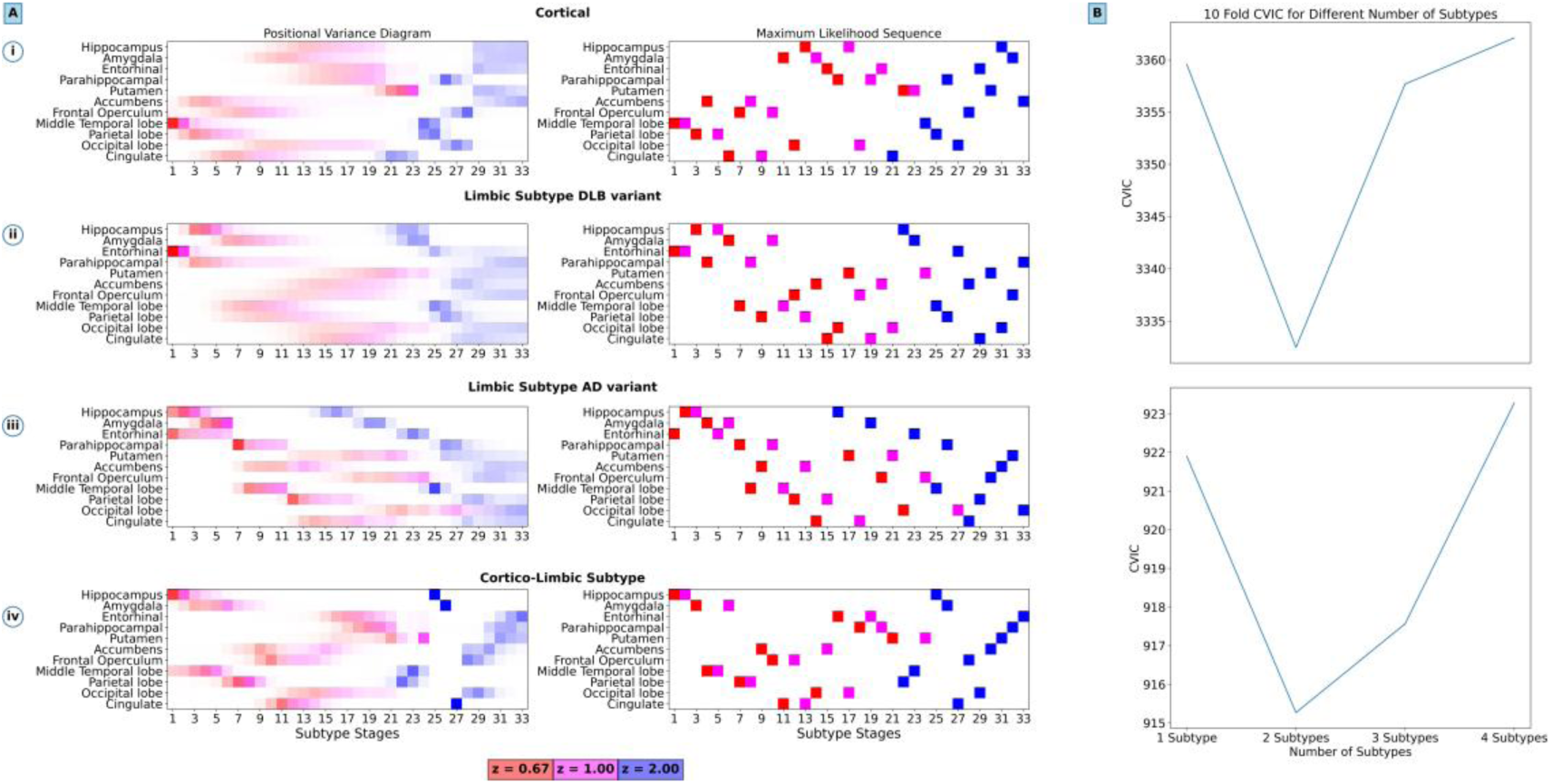
SuStaIn results for the DLB population and the AD population, and each population’s model validation. The figure represents each population’s subtypes: (i) Cortical and (ii) Limbic in the DLB population, and (iii) Limbic and (iv) Cortico-Limbic from the AD population. Each row contains three panels, from left to right: (A) Positional Variance Diagram displaying the uncertainty in the temporal evolution of regional atrophy (x-axis: stages 1-33; y-axis: brain regions), with intensity indicating probability of event (red: z-score=0.67, magenta: z-score=1, blue: z-score=2); (B) Maximum Likelihood Sequence showing the temporal evolution of regional atrophy, where colours indicate severity thresholds (same thresholds as before); (C) Cross-Validation Information Criterion for the model run for 1 through 4 subtypes, showing that 2 subtypes is the best fit.

Given the high degree of similarity with its homologue from the AD cohort and the strength of the relationship between their likelihoods, we discarded the Limbic subtype from the DLB variant, thus avoiding redundancy in the final transdiagnostic model. The other subtypes were combined – including the null subtype – into a final model (see ‘Transdiagnostic Subtyping Framework’) that we use for transdiagnostic subtyping and staging (differential diagnosis) of individuals from all cohorts.

### Data harmonization

Previous studies have shown that there are substantial ‘batch effects’ present in volumetric measurements derived from structural MRI scans, possibly confounding the underlying disease signal.^72^ We explore these effects on a subset of subjects where the original scans were available and use the MRIQC^73^ and NeuroHarmony^74^ tools to harmonize the data. We define the ‘batch effects’ as the scanner manufacturer, model, and field strength – all previously described to affect image quality and thus any measurement derived from the scan.^75^

We investigate the effect of scan manufacturer, model, and field strength in the biomarkers that the SuStaIn algorithm was trained on. This combination of ‘batch effects’ results in a total of 34 groups (detailed in **Table S1**). In this experiment, data is previously z-scored, and then NeuroHarmony model is trained on the groups that contain at least 20 samples and later applied to everyone. To understand the ‘batch effects’, we use Linear Discriminative Analysis (LDA) for visualization and the Lazy Classifier python package to explore a wide range of classifiers – the task being the correct assignment of the batch based on the set of biomarkers – and selected the top performers at any point and compared them. The LDA results show that there is, in fact, some ‘batch effect’ on the signal in some of the FreeSurfer outputs. Harmonization reduces the ‘batch effects’ (while preserving meaningful biological signal) as seen by the reduction in the clustering separation.

Nevertheless, the biomarkers selected as model inputs show no cluster separation, highlighting that LDA uses other metrics to discriminate between groups (**Figure S2**). In addition to this, the classification performance dropped after harmonization when compared to the non-harmonized data, but using the model-selected biomarkers also caused a drop in performance in the non-harmonized scenario, supporting the idea that these biomarkers retain little ‘batch effects’ (**Figure S3**). The results using SuStaIn show little difference. In both scenarios – harmonized and not harmonized data – the model converges to the two-subtype solution, with overlapping subtype-specific sequences, as shown by both the maximum likelihood sequences and the positional variance diagrams (**Figure S4**). Hellinger distance analysis shows strong similarity between harmonized and not harmonized pairs (0.137 and 0.078 for the Cortico-Limbic and Limbic subtypes). These results are in line with previous studies on SuStaIn where the use of harmonization techniques did not significantly change the model outputs.^55^

**Figure S2:**
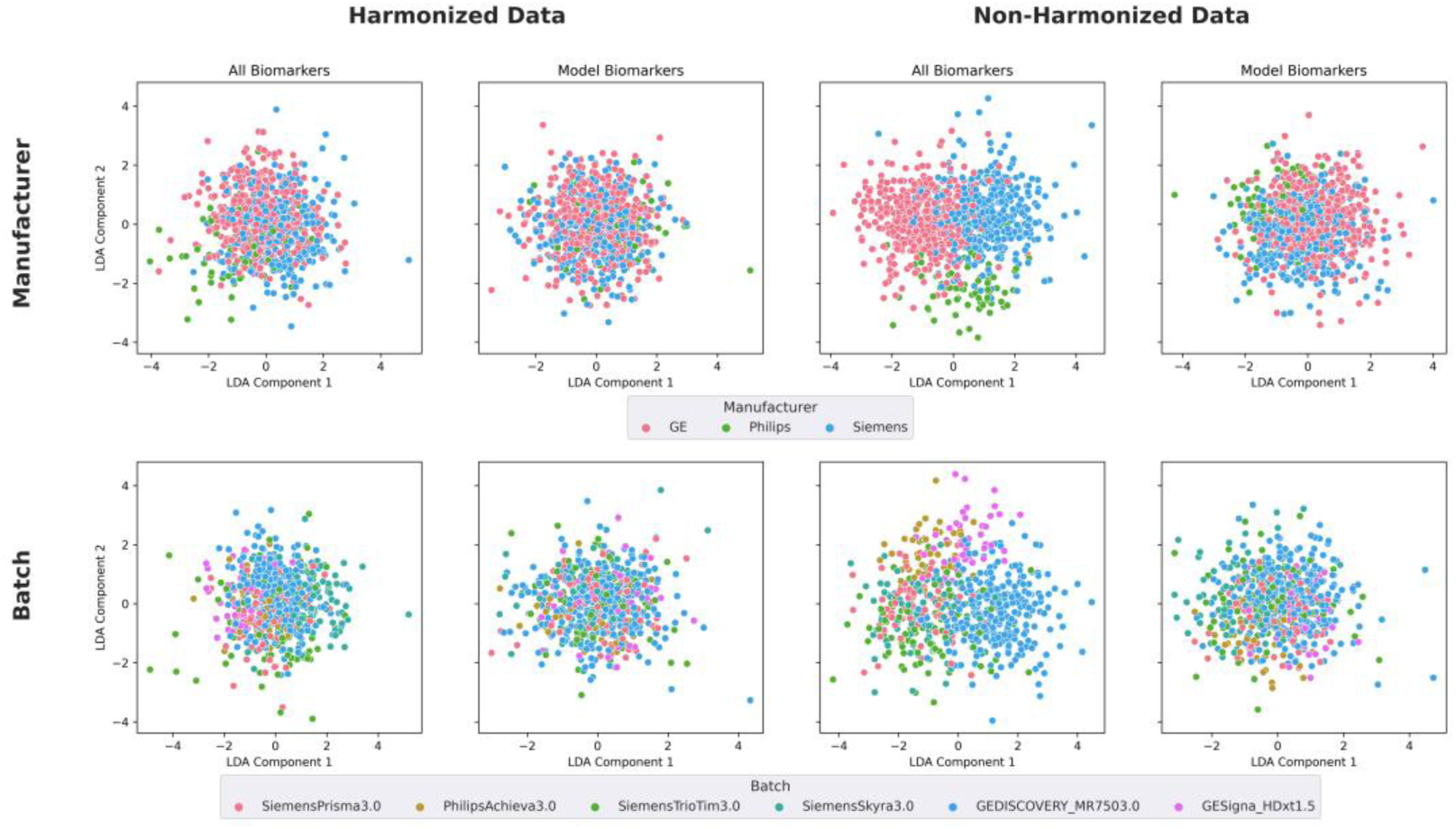
Linear discriminant analysis of neuroimaging data by manufacturer and batch (only top 6 with most subjects). Harmonized (left) and non-harmonized (right) data are shown with either all biomarkers or only model-selected biomarkers. Points are coloured by scanner manufacturer (top panels: GE; Philips; Siemens) or specific ‘batch’ (bottom panels).

**Figure S3:**
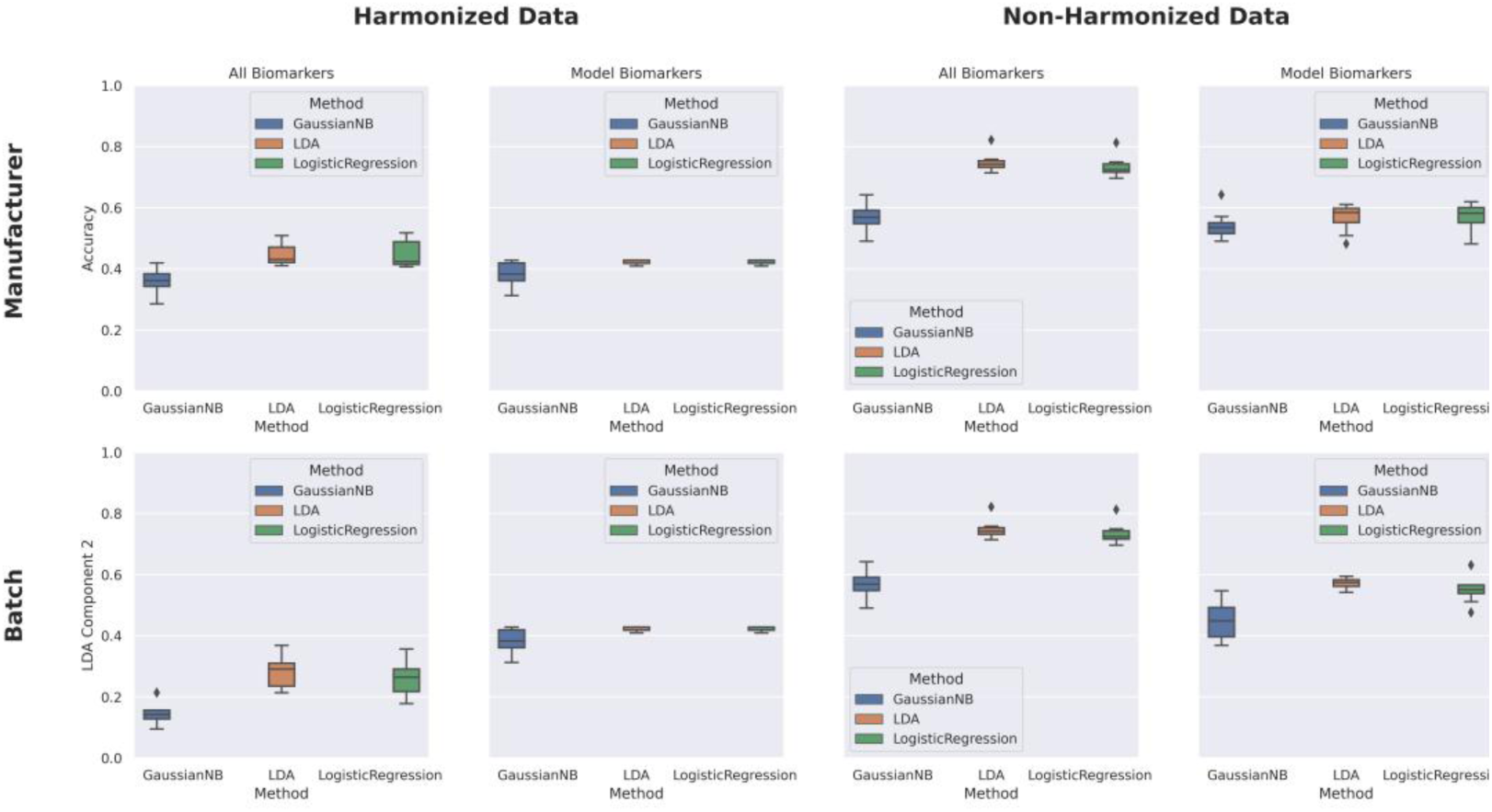
Classification accuracy using neuroimaging data of manufacturer and batch (only top 6 with most subjects). Harmonized (left) and non-harmonized (right) results are shown with either all biomarkers or only model-selected biomarkers. Boxplots are coloured by classification method (Gaussian Naive Bayes, blue; LDA, orange; Logistic Regression, green).

**Figure S4:**
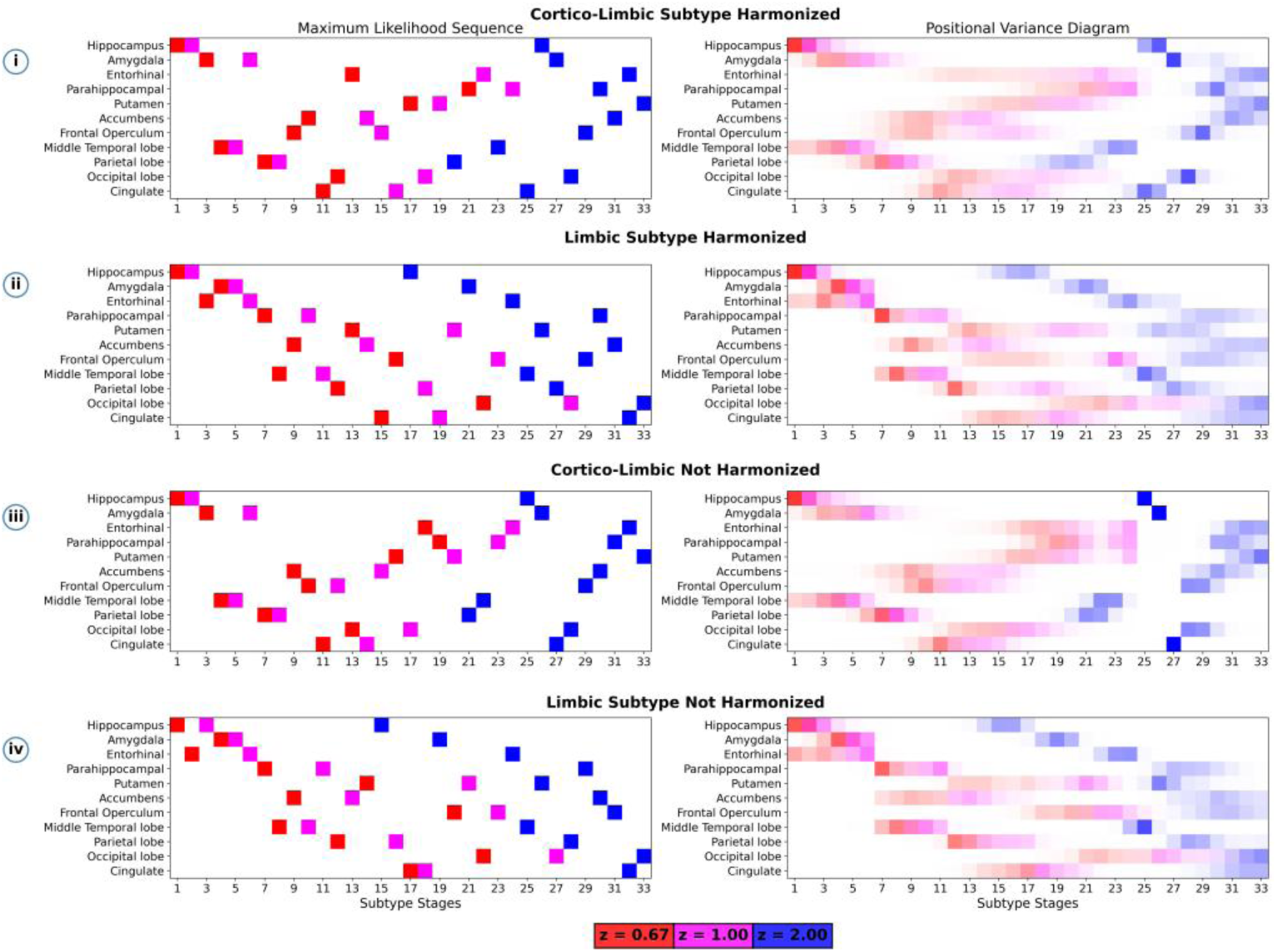
SuStaIn model results for AD using Harmonized and Not Harmonized data, showing subtype progression patterns. The figure is organized in four rows representing distinct subtypes: (i) Cortico-Limbic Harmonized, (ii) Limbic Harmonized, and (iii)Cortico-Limbic Not Harmonized, (iv) Limbic Not Harmonized. Each row contains three panels, from left to right: (A) Maximum Likelihood Sequence showing the temporal evolution of regional atrophy (x-axis: stages 1-33; y-axis: brain regions), where colours indicate severity thresholds (red: z-score=0.67, magenta: z-score=1, blue: z-score=2); (B) Positional Variance Diagram displaying the uncertainty in the ordering of events, with intensity indicating probability of event (same thresholds as before).

### The Null Subtype: Implementation and Implications

We test the null hypothesis that data from a given participant does not follow the model estimated clusters (these are the originally estimated clusters by SuStaIn) by adding the “Null” subtype to the model estimated ones, such that the data is now subtyped and staged across all. This Null subtype is characterized as a uniform probability distribution of a given event to happen for all biomarkers before any subsequent event happens for any biomarker (see **Figure S5** for the PVD).

**Figure S5:**
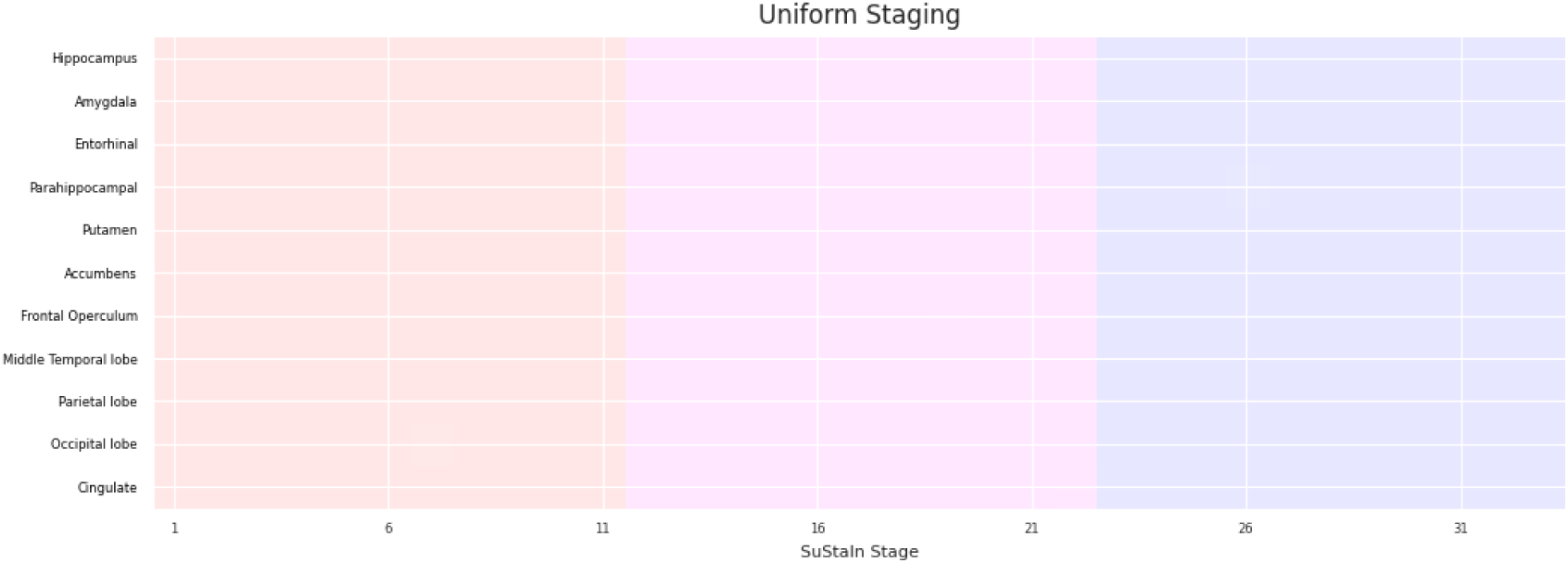
Positional Variance Diagram representation for the synthetic subtype included in the TranSuStaIn model framework to detect discordant data – that is, data that comes from a subtype trajectory different from one included in the model, or data corrupted by too much noise. For any given biomarker, the probability of the position of the first event is uniform across stages 1 to 11, for the second event is 12 to 22, and for the third event is 23 to 33.

A consequence of this framework, as opposed to the off-the-self (OTS) SuStaIn, is that fewer subjects are cross subtyped (that is, assigned to one subtype, and then a different one on a follow-up visit), especially across the Cortical and Limbic subtypes. In ADNI – where subjects have the most follow-up visits available (1567 unique subjects staged at stage 1 or above, >4 visits on average) – we observed that OTS SuStaIn would cross subtype 364 subjects in total (23.3%): 185 between Limbic and Cortico-Limbic, 165 between Cortico-Limbic and Cortical, and 74 between Cortical and Limbic – 30 subjects have gone through all three. Including the Null subtype reduces all numbers (there are now 1140 non-Null unique participants above stage 1), 168 cross subtyped (14.7%): 105 between Limbic and Cortico-Limbic, 64 between Cortico-Limbic and Cortical, and 5 between Cortical and Limbic – 3 subjects subtyped across all three.

Moreover, participants that are initially classified as Null will be subtyped as Cortical/Limbic/Cortico-Limbic in a subsequent visit as often as 35.8% out of those with at least two visits (N = 466), and 41% if we only account out of those with three or more visits (N = 385). This highlights that participants assigned to the Null, are (in this cohort) likely to be outliers due to noise in their data and thus the Null subtype assignment cannot be explored as a coherent group (in the way we have explored cognitive and biomarker differences across the other subtypes).

### Extended Analysis: Genetic and CSF including first non-Null visit

Some participants who were assigned to the Null Subtype at baseline, were assigned to other subtypes at later visits. In this extended analysis we added the first non-Null visit from these participants, which increased the sample size for our cross-sectional subtype comparisons. Our extended findings below reflect this through: a) confirming the same associations but with smaller p-values; and b) the new finding that amyloid status was associated with one subtype and not only explained by genetic differences.

We first re-assessed genetic risk prevalence across subtypes, where we added 146 participants (24 to the Cortical, 48 to the Cortico-Limbic, and 74 to the Limbic, see **Figure S6a**), to make a total of 935 participants. Prevalence of APOE epsiolon4 allele counts was again disproportionately distributed (χ^2^ = 41.7, P-value = 1.95E-08, V = 0.15), with carriers positively associated with Cortico-Limbic (residuals = 0.80 and 1.45, for 1 and 2 alleles respectively) and Limbic (residuals = 0.36 and 1.28, for 1 and 2 alleles respectively) subtypes. Participants assigned to the Cortical subtype were again positively associated with the absence of *APOE* ε4 alleles (residuals = 3.87).

We then re-assessed CSF status across subtypes, where we added 138 participants (24 to the Cortical, 32 to the Cortico-Limbic and 82 to the Limbic, see **Figure S6b**), to make a total of 541 participants. CSF status is still unevenly distributed across subtypes (χ^2^ = 59.1, P-value = 4.43E-12, V = 0.234), with A-/T-more prevalent in the Cortical subtype (residual = 4.91), A+/T-more prevalent in the limbic subtype (residual = 0.99), and A+/T+ more prevalent in the Cortico-Limbic subtype (residual = 2.97). Additionally, the Cortical subtype was strongly and negatively associated with A+/T+ status (residual = - 3.53) and the Cortico-Limbic with the A-/T-(residual = -3.37). The Limbic subtype had an almost expected assignment of A-/T- and A+/T+ participants, given the full cohort prevalence (residuals = -0.3 and -0.12, respectively).

**Figure S6:**
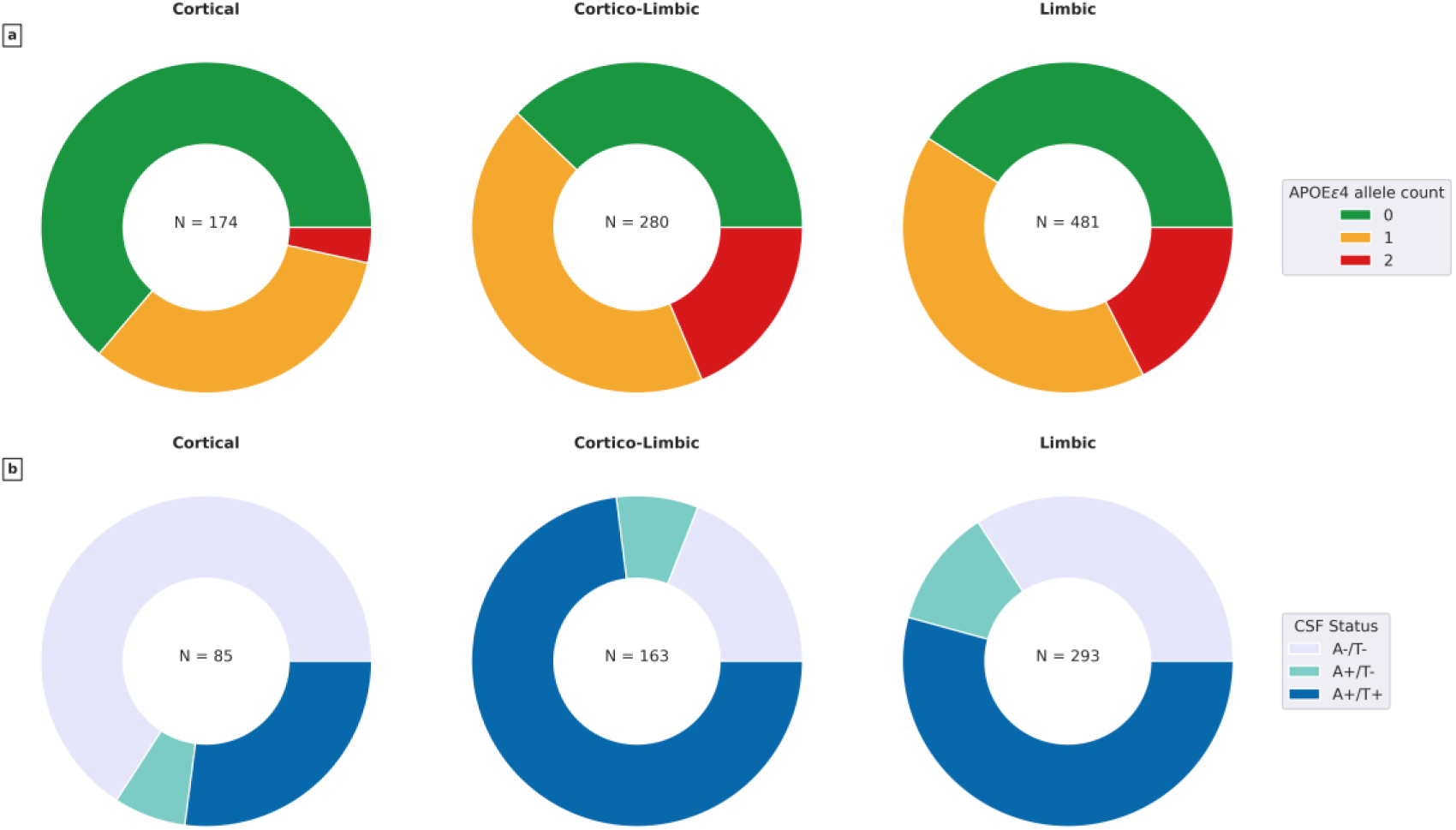
TranSuStaIn model subtype associations with genetic (panel a), AT(N) status (panel b) for the first non-Null visit of all participants. **a** Shows the proportion of *APOE* ε4 carriers in each subtype. **b** Shows the proportion positivity across Amyloid and Tau according to the CSF p-tau/Aβ ratio. A-/T-= Amyloid and Tau negative, A+/T-= Amyloid positive and Tau negative, A+/T+ = Amyloid and Tau positive.

We also re-assessed the CSF p-tau/Aβ ratio across subtypes and stages. When we do not correct for *APOE* ε4 allele count, we find stronger statistical differences across subtypes. The Cortical subtype has a lower expected ratio at early stages (ratio = 0.013, 99% CI = [0.005, 0.033]) compared to Cortico-Limbic (ratio = 0.026, 99% CI = [0.010, 0.066], P-value = 9.35E-05) and Limbic (ratio = 0.019, 99% CI = [0.007, 0.048], P-value = 1.42E-02, see **Figure S9**). Even when we correct for *APOE* ε4 allele count, we now find marginally significant differences across subtypes. Specifically, at early stages the Cortico-Limbic subtype exhibits a higher expected ratio (0.008, 99% CI = [0.004, 0.019], P-value = 1.53E-02) compared to the Cortical subtype (ratio = 0.006, 99% CI = [0.003, 0.013]) regardless of genetic risk.

Differences in SAA status (N = 463, 127 SAA+, 336 SAA-) were only apparent in the severity dimension for the Cortico-Limbic subtype. Using χ^2^ contingency test we found no association between SAA status and phenotype (P-value = 0.38, χ^2^ = 1.92, V = 0.065), with the prevalence of SAA+ across subtypes being 20.6%, 27.8%, and 28.9% for the Cortical, Cortico-Limbic and Limbic subtypes respectively. However, participants in the Cortico-Limbic subtype showed moderately significant and higher rate of SAA+ (37.73%) in the middle stages (> 11) when compared to those in the early stages (< 11, prevalence = 19.35%, P-value = 0.047, χ^2^ = 3.94, V = 0.185).

### PET biomarker profiles in early stages across subtypes

In ADNI patients with PET data available (<15% with Dementia), we compared Centiloid and Tau SUVR of brain lobes – temporal, occipital, parietal, and frontal – across subtypes and with Stage Zero individuals. These participants were largely assigned to the Stage Zero group (62.3%), and if subtyped, they showed moderate brain atrophy – most participants (75 %) were assigned to stage 12 or prior. We grouped this early atrophy participants into their subtypes and assessed group-level comparisons.

We found significantly different Centiloids level (Krustal-Wallis test P-value < 9.79E-19, see **Figure S7**) across participants from different groups. Participants from both Limbic and Cortico-Limbic subtypes (N = 180 and 71, respectively) showed elevated Centiloids (mean value = 53.99, 99% CI = [44.29, 63.69]; and mean value = 60.85, 99% CI = [43.69, 78.00], respectively) when compared to those assigned to Stage Zero (N = 732, mean value = 23.49, 99% CI = [19.86, 27.11], P-value < 7.41E-09). On the other hand, we did not find differences between Stage Zero and Cortical subtype participants (N = 56, mean value = 20.95, 99% CI = [5.25, 36.65], P-value = 0.141). These results show a trend of elevated Aβ deposition in participants assigned to both Limbic and Cortico-Limbic subtypes.

We also found significantly different levels of Tau PET SUVR in all brain lobes (P-value < 2.58E-03, see **Figure S8**) across groups. We found that participants assigned to the Cortical subtype had similar SUVR to those assigned to Stage Zero (N = 18 and 448 participants per group, respectively; Mann-Whitney U test P-value > 0.05) for all brain lobes. On the other hand, participants assigned to the Cortico-Limbic subtype (N = 20) had significantly higher SUVR than those assigned to the Stage Zero (P-value < 1.19E-02, Cliff’s delta > 0.33) for all brain regions except the occipital lobe (P-value = 0.052, Cliff’s delta = 0.257). Participants assigned to the Limbic subtype (N = 99) had statistically higher SUVR than Stage Zero participants in all lobes (P-value < 1.48E-02, Cliff’s delta > 0.2). In the Limbic and Cortico-Limbic subtypes, we found that Tau deposition in the brain overlapped with the cortical regions in the brain that are affected earlier in their respective subtype-specific progression patterns. However, participants in the Cortical subtype showed no Tau deposition despite having moderate cortical atrophy.

**Figure S7:**
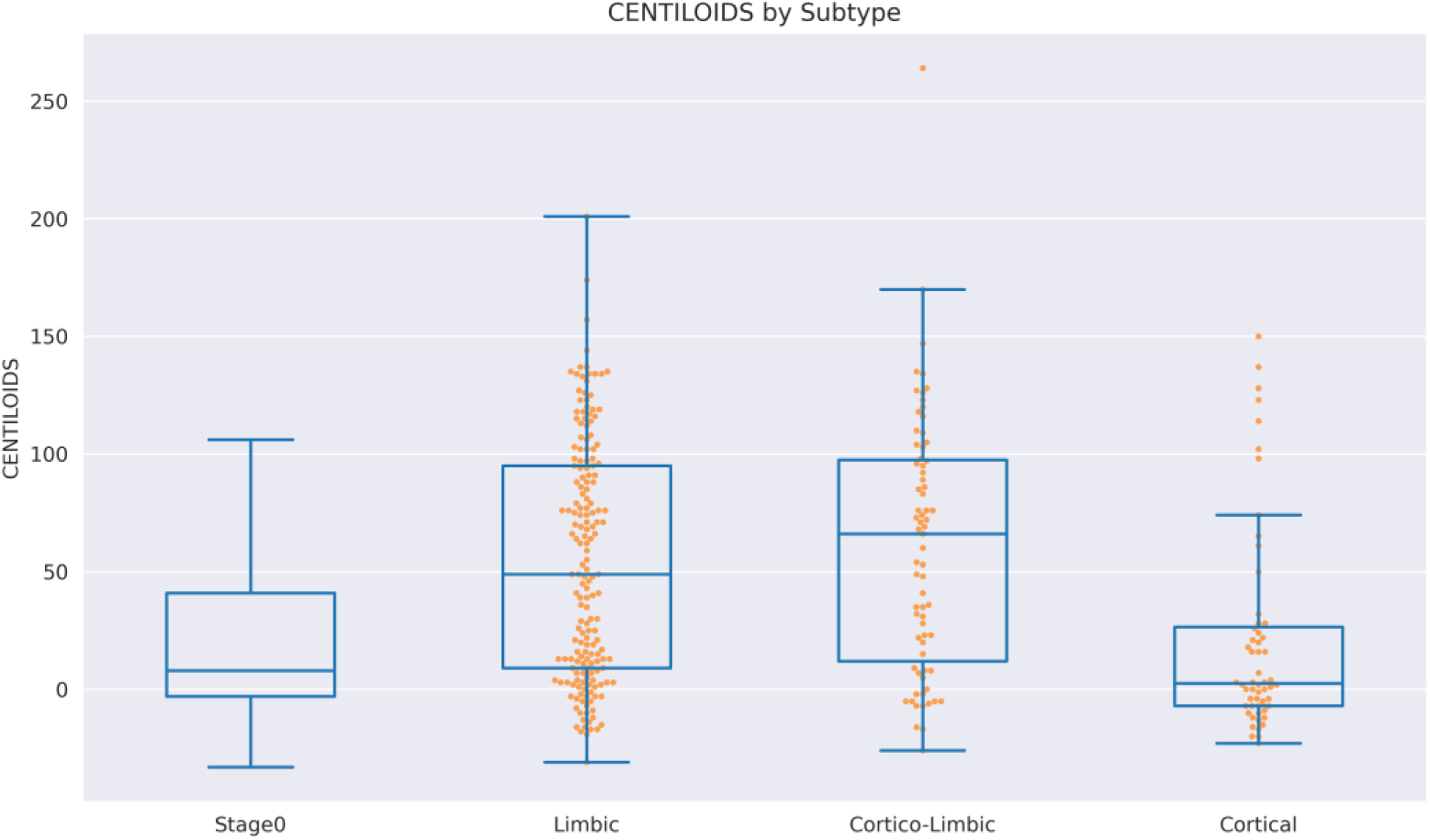
Amyloid PET Centiloid value distributions of participants assigned to each subtype, and Stage Zero (sub-threshold atrophy). Participants assigned to Stage Zero and Cortical subtype show the least amount of amyloid deposition on PET scan, while those with the Limbic or Cortico-Limbic pattern display a higher trend.

**Figure S8:**
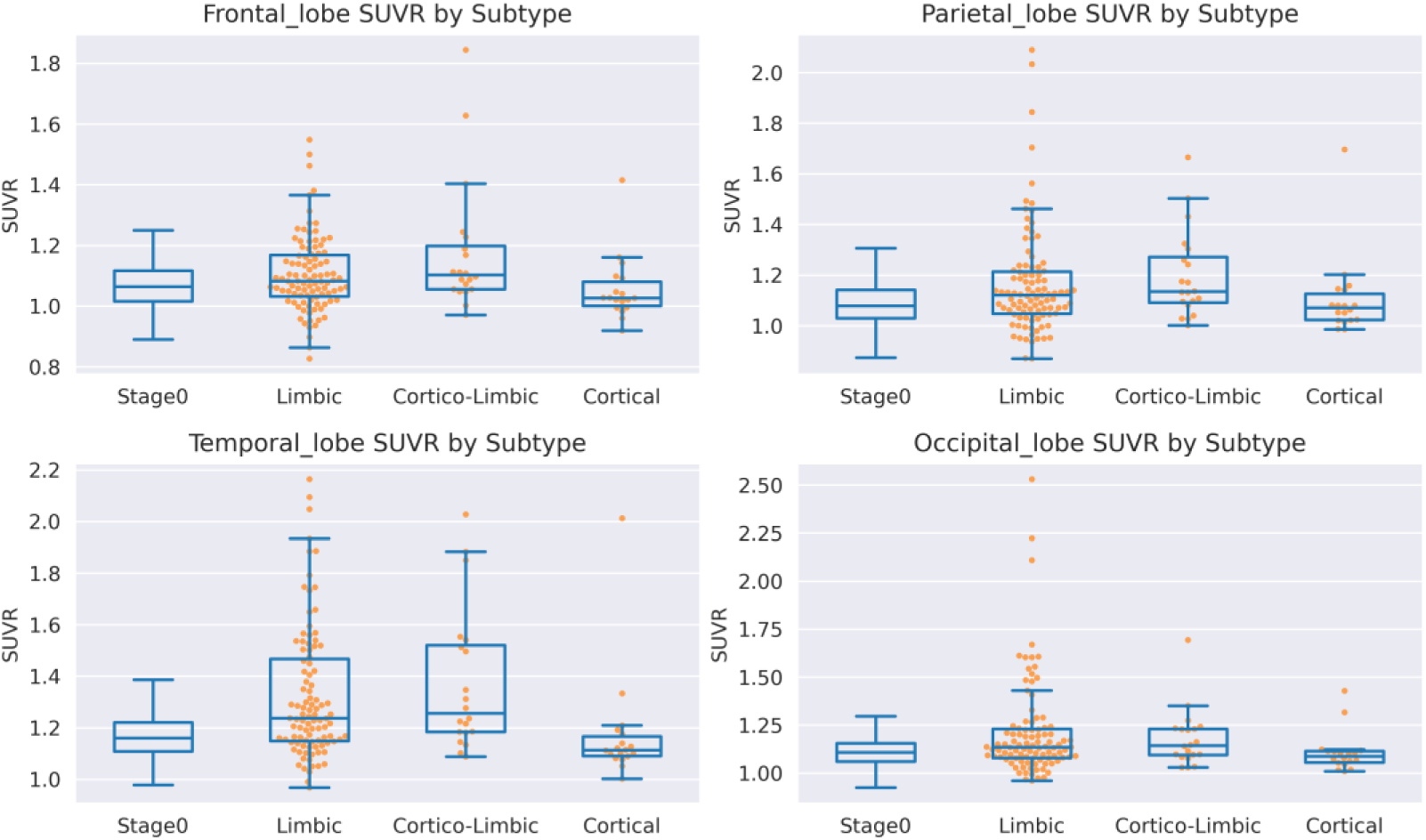
Tau PET SUVR distributions per brain lobe of participants assigned to each Subtype, and Stage Zero (sub-threshold atrophy). Participants assigned to the Stage Zero or the Cortical subtype display similarly low SUVR values across all regions, while those assigned to the Limbic and Cortico-Limbic subtypes show higher values, particularly for the frontal, parietal, and temporal lobes.

**Table S1:** Imaging derived phenotypes used as the model input features. Regions of interest and their component sub-regions are shown, from the Desikan-Killiany atlas. ROI = Region of Interest

| ROI | Desikan-Killiany parcellation. |
| --- | --- |
| Hippocampus | Left and right hippocampus. |
| Amygdala | Left and right amygdala. |
| Entorhinal | Left and right entorhinal. |
| Parahippocampal | Left and right parahippocampal. |
| Accumbens | Left and right accumbens. |
| Putamen | Left and right putamen. |
| Frontal Operculum | Left and right pars opercularis, pars orbitalis, and pars triangularis. |
| Parietal Lobe | Left and right inferior parietal, postcentral, precuneus, superior parietal, and supramarginal. |
| Middle Temporal | Left and right middletemporal. |
| Occipital Lobe | Left and right cuneus, lateral occipital, lingual, and pericalcarine. |
| Cingulate | Left and right, caudal anterior cingulate, isthmus cingulate, posterior cingulate, rostral anterior cingulate. |

**Table S2:** Demographics of subjects by Dataset. CU = Cognitively Unimpaired; MCI = Mild Cognitive Impairment; Dem = Dementia; STD = Standard Deviation; MMSE = Mini Mental State Examination; MoCA = Montreal Cognitive Assessment; AD = Alzheimer’s disease; LBD = Lewy body disease; DLB = Dementia with Lewy bodies.

|  | ADNI |  |  | NACC |  |  | PDBP |
| --- | --- | --- | --- | --- | --- | --- | --- |
|  | CU | MCI | Dem | CU | MCI | Dem | Dem |
| Sample | 827 | 981 | 360 | 875 | 566 | 536 | 16 |
| Age | 72.43 | 73.00 | 74.53 | 71.33 | 72.98 | 72.73 | 72.45 |
| (± std) | (± 6.60) | (± 7.48) | (± 7.82) | (± 9.62) | (± 8.72) | (± 9.82) |  |
|  |  |  |  |  |  |  | (±<br>8.93) |
| Male (Female) | 374<br>(453) | 579 (402) | 198 (162) | 313<br>(562) | 284<br>(282) | 265<br>(271) | 16 (0) |
| MMSE<br>(± std) | 29.05<br>(± 1.15) | 27.61<br>(± 1.88) | 23.13<br>(± 2.32) | 29.18<br>(± 1.09) | 26.90<br>(± 2.45) | 21.08<br>(± 5.85) | NA |
| MoCA<br>(± std) | 26.00<br>(± 2.52) | 23.21<br>(± 3.20) | 17.09<br>(± 4.57) | 26.79<br>(± 2.35) | 22.29<br>(± 3.32) | 15.25<br>(± 5.55) | 19.94<br>(±<br>6.09) |
| Cognitive Domains |  |  |  |  |  |  |  |
| Memory | N = 772<br>0.83<br>(± 0.47) | N = 925<br>0.10<br>(± 0.52) | N = 337<br>-0.78<br>(± 0.35) | N = 789<br>1.09<br>(± 0.58) | N = 527<br>0.10<br>(± 0.58) | N = 438<br>-0.87<br>(± 0.68) | NA |
| Executive | N = 770<br>0.75<br>(± 0.47) | N = 924<br>0.32<br>(± 0.55) | N = 336<br>-0.41<br>(± 0.67) | N = 788<br>0.68<br>(± 0.59) | N = 526<br>0.02<br>(± 0.63) | N = 402<br>-0.66<br>(± 0.71) | NA |
| Language | N = 771<br>0.82<br>(± 0.48) | N = 925<br>0.38<br>(±0.50) | N = 341<br>-0.20<br>(± 0.58) | N = 792<br>0.97<br>(± 0.57) | N = 528<br>0.28<br>(± 0.56) | N = 433<br>-0.52<br>(0.67) | NA |
| Visuospatial | N = 345<br>0.127<br>(± 0.29) | N = 564<br>0.01<br>(± 0.37) | N = 269<br>-0.24<br>(± 0.59) | NA | NA | NA | NA |
| CSF Proteins [pg/mL] |  |  |  |  |  |  |  |
| Amyloid β | N = 248<br>1045.97<br>(±<br>383.55) | N = 470<br>837.30<br>(± 342.26) | N = 189<br>628.55<br>(±<br>239.92) | NA | NA | NA | NA |
| Phospho-Tau | N = 343<br>21.69<br>(± 8.94) | N = 551<br>28.25<br>(± 14.85) | N = 197<br>37.57<br>(± 15.57) | NA | NA | NA | NA |
| ApoE ε4 |  |  |  |  |  |  |  |
| 0 | 521 | 470 | 109 | NA | NA | NA | NA |
| 1 | 221 | 346 | 162 | NA | NA | NA | NA |
| 2 | 21 | 104 | 74 | NA | NA | NA | NA |
| DLB Core Features (Yes/No) |  |  |  |  |  |  |  |
| Hallucinations | NA | NA | NA | 0/6 | 4/26 | 23/46 | NA |
| Cognitive Fluctuations | NA | NA | NA | 0/1 | 2/11 | 13/22 | NA |
| REM Disorder | NA | NA | NA | NA | 4/9 | 10/8 | NA |
| Parkinsonism | NA | NA | NA | 2/3 | 5/10 | 11/31 | NA |
| Neuropathology |  |  |  |  |  |  |  |
| AD | 9 | 28 | 19 | 16 | 21 | 117 | NA |
| LBD | 2 | 12 | 2 | 2 | 9 | 10 | NA |
| AD + LBD | 0 | 9 | 16 | 6 | 12 | 47 | NA |
| Amyloid PET |  |  |  |  |  |  |  |
| Centiloids | N = 295<br>19.03<br>(± 31.63) | N = 277<br>36.59<br>(± 46.63) | N = 54<br>86.83<br>(± 50.21) | NA | NA | NA | NA |
| Tau PET | N = 268 | N = 158 | N = 54 |  |  |  |  |
| Left Temporal Lobe SUVR | 1.16<br>(± 0.10) | 1.31<br>(± 0.33) | 1.61<br>(± 0.43) | NA | NA | NA | NA |
| Right Temporal Lobe SUVR | 1.16<br>(± 0.10) | 1.30<br>(± 0.30) | 1.57<br>(± 0.41) | NA | NA | NA | NA |
| Left Occipital Lobe SUVR | 1.10<br>(± 0.07) | 1.18<br>(± 0.23) | 1.46<br>(± 0.56) | NA | NA | NA | NA |
| Right Occipital Lobe SUVR | 1.10<br>(± 0.07) | 1.19<br>(± 0.29) | 1.42<br>(± 0.53) | NA | NA | NA | NA |
| Left Parietal Lobe SUVR | 1.07<br>(± 0.08) | 1.15<br>(± 0.23) | 1.42<br>(± 0.52) | NA | NA | NA | NA |
| Right Parietal Lobe SUVR | 1.07<br>(± 0.09) | 1.14<br>(± 0.23) | 1.40<br>(± 0.53) | NA | NA | NA | NA |
| Left Frontal Lobe SUVR | 1.04<br>(± 0.09) | 1.08<br>(± 0.17) | 1.29<br>(± 0.39) | NA | NA | NA | NA |
| Right Frontal Lobe SUVR | 1.04<br>(± 0.09) | 1.09<br>(± 0.17) | 1.29<br>(± 0.36) | NA | NA | NA | NA |
|  | E-DLB |  |  | ViPD | USyd |  |  |
|  | CU | MCI | Dem | Dem | CU | Dem |  |
| Sample | 189 | 129 | 281 | 42 | 32 | 21 |  |
| Age<br>(± std) | 67.12<br>(± 9.21) | 72.91<br>(± 6.74) | 75.20<br>(± 8.21) | 72.43<br>(± 5.53) | 66.58<br>(± 7.82) | 74.49<br>(± 6.35) |  |
| Male (Female) | 96 (93) | 81 (48) | 170 (111) | 38 (4) | 19 (13) | 18 (3) |  |
| MMSE<br>(± std) | 28.47<br>(± 2.81) | 26.22<br>(± 2.60) | 21.78<br>(± 5.01) | 24.86<br>(± 3.98) | NA | 22.7<br>(± 5.69) |  |
| MoCA | 24.52 | 16.38 | 15.38 | 20.93 | 28.11 | 18.7 |  |

| (± std) | (± 5.55) | (± 7.52) | (± 7.52) | (± 5.63) | (± 2.04) | (± 6.02) |
| --- | --- | --- | --- | --- | --- | --- |
| DLB Core Features<br>(Yes/No) |  |  |  |  |  |  |
| Hallucinations | 1/53 | 33/91 | 127/129 | 30/8 | NA | 10/10 |
| Cognitive Fluctuations | 1/31 | 47/42 | 131/105 | 33/5 | NA | 15/5 |
| REM Disorder | 0/33 | 73/51 | 116/88 | 29/9 | NA | 20/1 |
| Parkinsonism | 1/0 | 36/43 | 97/112 | 31/7 | NA | 16/5 |

**Table S3:**
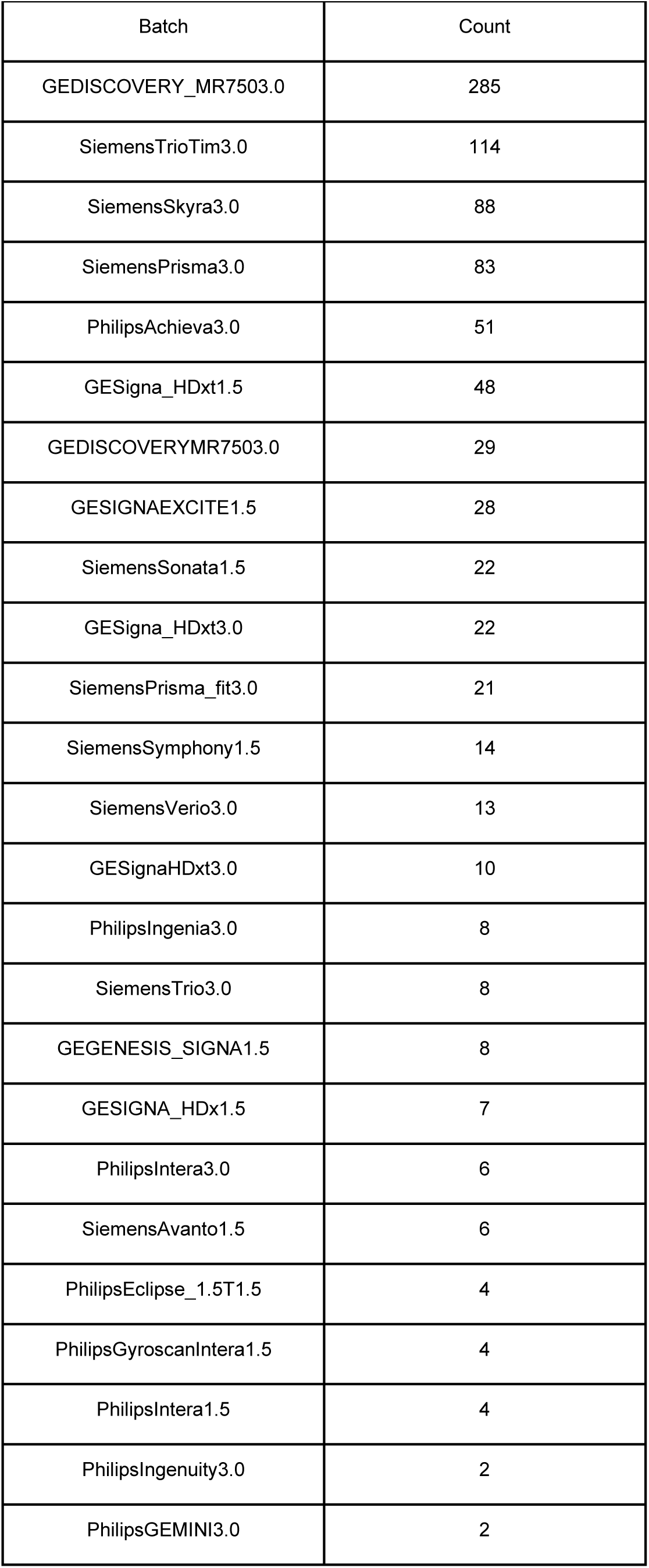
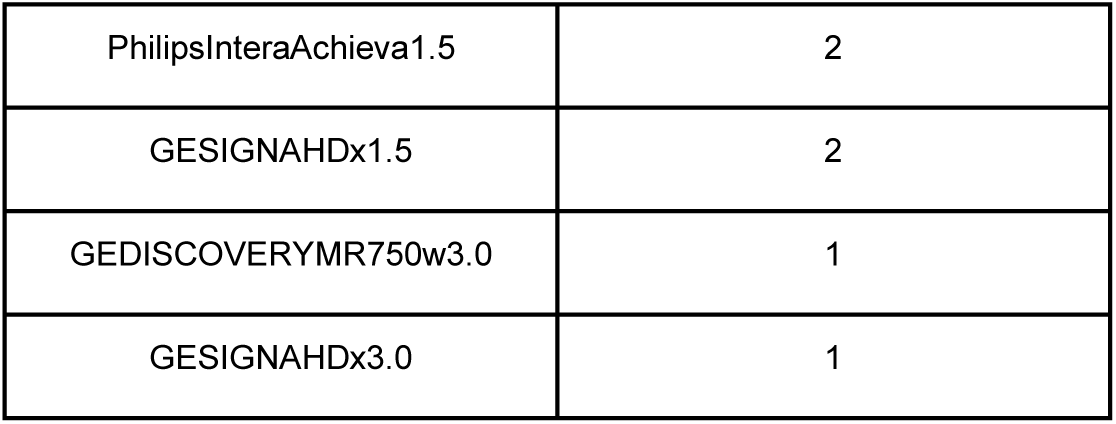
Batch effects (as manufacturer, model and field strength) groups and counts of samples.

**Figure S9:**
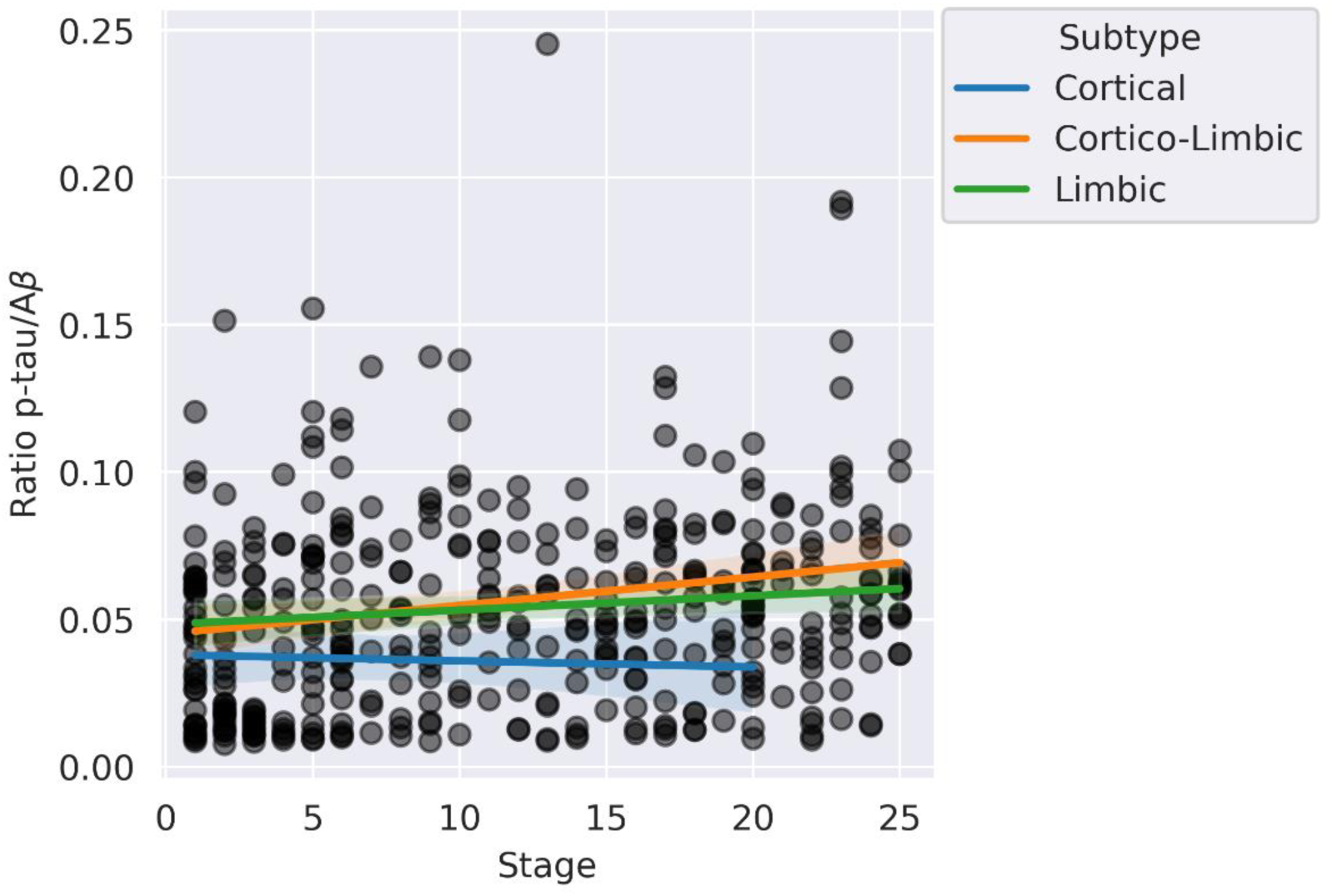
TranSuStaIn subtype and stage assignments associations with CSF p-tau/Aβ ratio. CSF = Cerebrospinal fluid, p-tau = phosphorylated tau, Aβ = amyloid-β.

**Figure S10:**
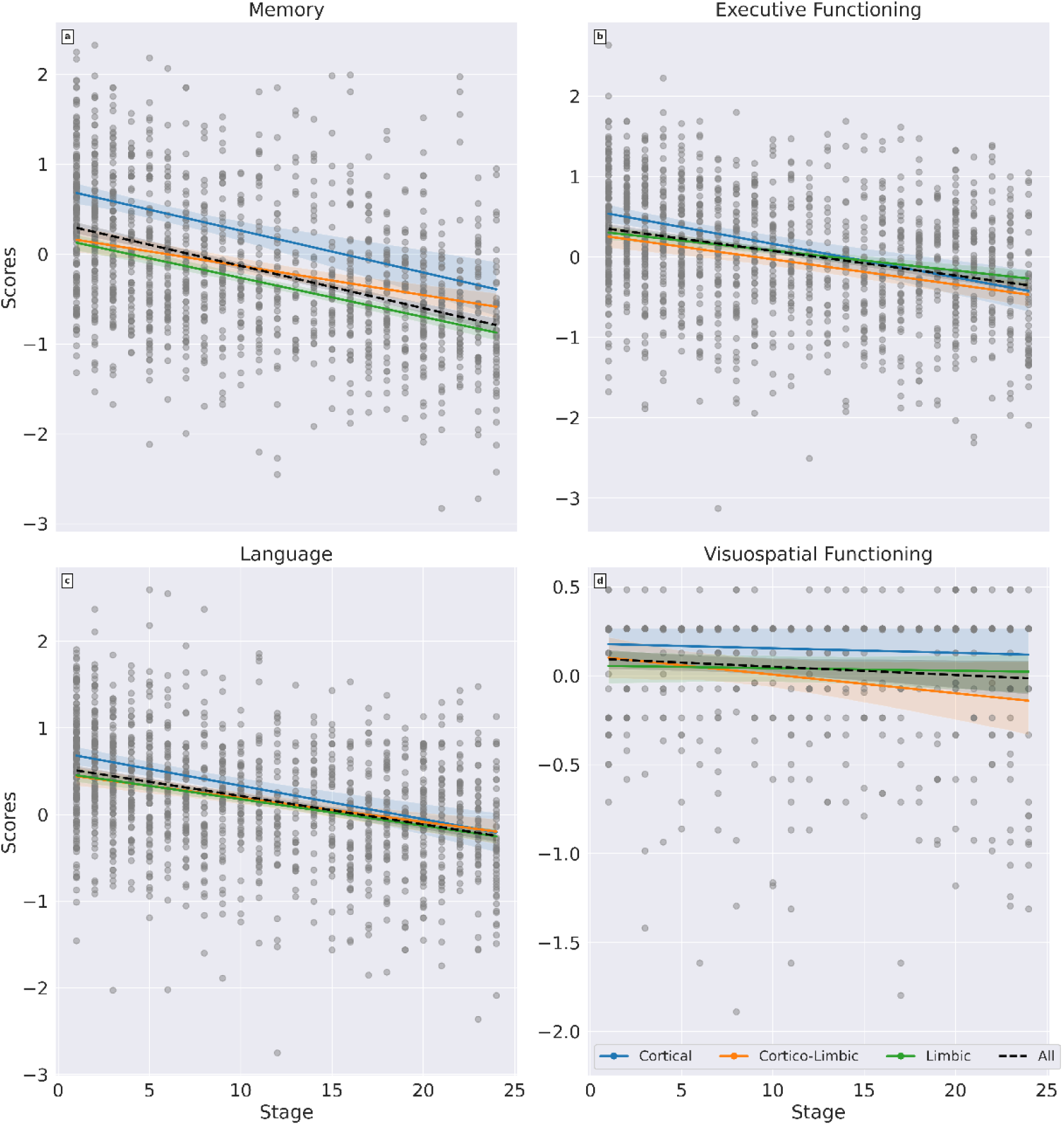
TranSuStaIn subtype and stage assignments associations with domain-specific cognitive scores – memory (a), executive (b), language (c), and visuospatial (d). These scores are harmonized across different cohorts by ADSP-PHC,^35^ and available for a subset of ADNI and NACC participants in our study.

## Notes

### Author Declarations

UCL Research Ethics Committee UCL Department of Computer Science Research Ethics Committee

### Summary of Updates

new participants included from the EDLB consortium and data from alpha-synuclein CSF, amyloid and tau PET analysis.

